# Sub-cellular level resolution of common genetic variation in the photoreceptor layer identifies continuum between rare disease and common variation

**DOI:** 10.1101/2022.03.15.22272407

**Authors:** Hannah Currant, Tomas W Fitzgerald, Praveen J Patel, Anthony P Khawaja, UK Biobank Eye and Vision Consortium, Andrew R Webster, Omar A Mahroo, Ewan Birney

**Affiliations:** European Molecular Biology Laboratory, European Bioinformatics Institute, Cambridge, UK; Novo Nordisk Foundation Center for Protein Research, Faculty of Health and Medical Sciences, University of Copenhagen, Copenhagen, Denmark; NIHR Biomedical Research Centre, Moorfields Eye Hospital NHS Foundation Trust UCL Institute of Ophthalmology, London, UK; Section of Ophthalmology, King’s College London, St Thomas’ Hospital Campus, London, UK; Physiology, Development and Neuroscience, University of Cambridge, Cambridge, UK

## Abstract

Photoreceptor cells (PRCs) are the light-detecting cells of the retina. Such cells can be non-invasively imaged using optical coherence tomography (OCT) which is used in clinical settings to diagnose and monitor ocular diseases. Here we present the largest genome-wide association study of PRC morphology to date utilising quantitative phenotypes extracted from OCT images within the UK Biobank. We discovered 111 loci associated with the thickness of one or more of the PRC layers, many of which had prior associations to ocular phenotypes and pathologies. We further identified 10 genes associated with PRC thickness through gene burden testing using exome data. In both cases there was a significant enrichment for genes involved in rare eye pathologies, in particular retinitis pigmentosa. There was evidence for an interaction effect between common genetic variants, *VSX2* involved in eye development and *PRPH2* known to be involved in retinal dystrophies. We further identified a number of genetic variants with a differential effect across the macular spatial field. Our results suggest a continuum between normal and rare variation which impacts retinal structure, sometimes leading to disease.

## Introduction

The retina is a layered structure at the back of the eye, responsible for receiving light stimulus and converting it to neurological signal which is interpreted by the visual system of the brain. Each retinal layer comprises different cells that are responsible for particular stages of the signal conversion process. The central area of the retina is termed the macula, and has a valley like structure. At the centre of the macula, the bottom of the valley structure, is the fovea which is the area of the retina responsible for highest acuity vision [1]. The retina can be imaged using optical coherence tomography (OCT). OCT produces high resolution, three-dimensional representations of the retina in which the different retinal layers and macular structure can be identified (fig. 1A). OCT is a non-invasive imaging method commonly used in the clinic to diagnose and monitor a number of ophthalmic conditions [2].

**Fig 1.**
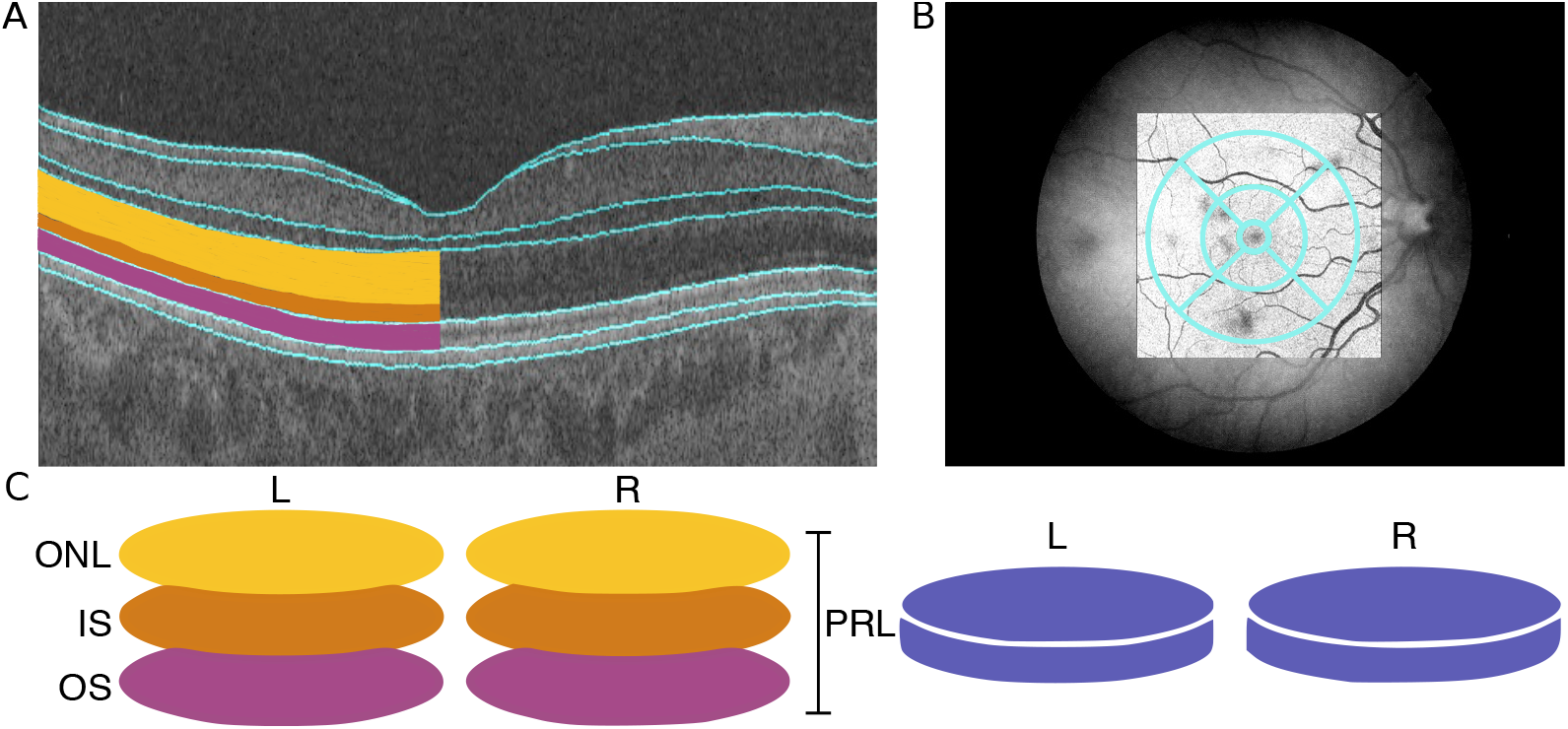
Data available from OCT images to describe morphology of the photoreceptor cell layer. (A) An optical coherence tomography (OCT) image of the cross-section of the retina. The layers are segmented, outlined in blue, by the Topcon advanced boundary software (TABS). On the left-hand side of the image, the PRC layers have been shaded: The outer nuclear layer (ONL) in yellow, the inner segment (IS) in orange and the outer segment (OS). (B) The early treatment of diabetic retinopathy study (ETDRS) segmentation grid super imposed on a fundus image of the retina. The ETDRS grid is composed of nine segments arranged in a bulls-eye. (C) A schematic of the data utilised to describe PRC morphology as an input phenotype in genome-wide association studies (GWAS). The ONL, IS and OS, combined, comprise the photoreceptor layer (PRL).

The photoreceptor cells (PRCs) are found towards the back of the eye within the retina, backed by the retinal pigment epithelium layer (RPE) (fig. 1A), and are responsible for detecting light photons and generating electrical response through the process of phototransduction. They consist of two main types, the rods and cones. Rods are responsible for vision in low light but offer lower visual acuity and lack colour definition. Cones are active in higher light levels, and confer colour and high acuity vision. The cones are densely populated at the centre of the retina, the fovea, where there are no rods. Conversely, rods are most densely populated more peripherally, where cone denisty is low. The PRC layer can be further divided into three component intracellular layers; the outer nuclear layer (ONL), which comprises the cell body of the PRC, including the cell nucleus; the inner segment (IS), which contains both the PRC’s mitochondria and ribosomes required for photopigment assembly; and the outer segment (OS), which interfaces with the RPE and contains the stacked membranes in which photopigments are stored.

There are numerous pathologies, both rare and common, that affect the morphology of the PRCs. Notably, age-related macular degeneration (AMD) is defined by the accumulation of lipid deposits, known as drusen, under the RPE. This causes damage to the RPE and subsequently degradation of the PRCs [3]. AMD causes progressive and irreversible loss of visual function, and is a major cause of blindness in older people. [4].

Many retinal dystrophies affect the photoreceptors [5]. These rare inherited disorders are due to highly penetrant mutations in genes expressed in PRCs or less commonly the RPE, resulting in retinal dysfunction with or without degeneration of the outer retinal layers. These can affect predominantly the central retina (macular dystrophies), the rod photoreceptors (rod-cone dystrophy or retinitis pigmentosa), or the cone photoreceptors (cone and cone-rod dystrophies). Though individually rare, taken together these diseases are a major cause of blindness in children and working age adults.

There have been previous studies on retinal morphology with known associations to age, sex, and ethnicity [6, 7]. Previous studies have further looked at the genetic variation underlying the overall retinal thickness [8] and the thickness of the inner retinal layers [9]. These studies identified loci with prior associations to general retinal development and ophthalmic disease.

In this paper, we utilise data within the UK Biobank to interrogate how genetic variation affects PRC morphology using quantitative phenotypes extracted from OCT images. The UK Biobank provides a large-scale rich resource of both genotypic and phenotypic data. Here we conduct genome-wide association studies (GWAS) of the thickness of the component PRC layers, the ONL, IS and OS. Following meta analysis of the three PRC layers, 111 loci are significantly associated with the thickness of one or more PRC layers. These loci include variants with prior associations to ophthalmic phenotypes, including ophthalmic diseases. We further explored the genetic variation of the morphology of the retina at a higher resolution by considering the retinal layer thickness in different concentric areas of the macula. This identified a number of loci that appear to differentially affect PRC morphology across the retinal field. Interestingly, several of these loci have prior associations to retinal diseases whose pathological action bears similarity with the spatial variation in genetic effect on morphology. Further analysis utilising the exome sequence data identified 10 genes associated with the thickness of one or more of the PRC layers. Several of these genes also had prior associations to ocular phenotypes. Additional analysis identified interaction between two of the common genetic variants, *VSX2* and *PRPH2*, affecting outer retinal thickness. We further explored the spatial genetic effect of our discovered genetic variants to capitalise on more of the available dimensionality of the phenotype data.

## Methods

### Ethics Statement

The UK Biobank study was conducted with the approval of the North-West Research Ethics Committee (ref 06/MRE08/65), in accordance with the principles of the Declaration of Helsinki, and all participants gave written informed consent. The research presented here has been conducted using the UK Biobank Resource under Application Number 2112.

### UK Biobank Data

The UK Biobank is a cohort comprising ∼500,000 individuals recruited through the NHS registry at age 40-69 from across the UK. Individuals were were not selected on the basis of having disease, resulting in a broad cross-section of the UK population. For all participants, a number of baseline physical measurements were taken and an extensive questionnaire completed comprising information on lifestyle, demographic and socioeconomic factors. Additionally, blood and urine samples were collected and further tests including a heel-bone ultrasound, bio-impedance, hand-grip strength, spirometry, blood pressure and several cognitive tests were performed. Each individual was further genotyped and exome sequenced. The participants also agreed to ongoing linkage of their medical records [10].

### Ophthalmic Measurements and Optical Coherence Tomography

A subset of 132,041 participants completed an eye examination. The measures collected included best corrected visual acuity using a logarithm of the minimum angle of resolution (logMAR) chart (Precision Vision, LaSalle, Illinois, USA). Visual acuity was measured with participants wearing their distance glasses at 4m, or at 1m if a participant was unable to read letters at 4m. Participants were asked to read from the top of the chart downwards, with the test terminated when two or more letters were read incorrectly. Further, refractive error was measured using a Tomey RC-5000 auto refraktometer (Tomey Corp., Nagoya, Japan) [11]. For each eye, up to 10 measurements were taken and the most reliable measure was automatically recorded. Intraocular pressure (IOP) was measured using an Reichert Ocular response Analyzer [12] from which corneal compensated IOP was calculated that accounts for rigidity of the cornea [13].

#### Optical Coherence Tomography Imaging Data

A further subset of 67,321 individuals underwent Spectral Domain Optical Coherence Tomography (SD-OCT) imaging. OCT imaging was conducted using the the Topcon 3D OCT1000 Mark II machine using 3-dimensional 6 × 6mm^2^ macular volume scan mode (512 A scans per B scan; 128 horizontal B scans). The imaging was completed on undilated eyes in a dark room following other eye measurements [13]. The right eye was consistently imaged first, and the majority of individuals had imaging repeated in their left eyes [12].

The images were processed using the industry standard Topcon advanced boundary software (TABS) [14]. This software performs automated retinal layer segmentation. It calculated thickness values for each of the retinal layers averaged across the macular field, as well as in each of the subfields of the early treatment of diabetic retinopathy study (ETDRS) grid [15]. The ETDRS is a nine-segment grid arranged as a bullseye and is commonly used during clinical assessment of the macula including the PRCs (fig. 1B).

### Quality Control of Genotypic and Phenotypic Data

Participants who were recommended for exclusion from genetic studies by the UK Biobank were removed from the dataset. The most densely populated well-mixed population was selected for using principal component analysis. Individuals within a defined euclidean distance of the mean of the selected population, as identified by comparison to the HapMap Phase III study [16] in PC1-PC2 space were selected. Individuals were further excluded if they were related to third degree or more [17].

In addition to the genotypic quality control, rigorous quality control was applied to the phenotypic data. Exclusion/inclusion criteria were applied to the OCT images and the quantitative measures derived from them utilising methods previously implemented in Patel *et al*. (2016) [18]. In line with this method all participants with an OCT image quality score *<*45 were removed from the study. Further, individuals with values within the poorest 20% of the population in each of the OCT segmentation indicators were removed. These segmentation indicators include: Inner limiting membrane (ILM) indicator, a measure of the minimum localised edge strength around the ILM boundary across the entire OCT scan. ILM indicator is indicative of blinks, severe signal fading, and segmentation errors; Valid count, used to identify significant clipping in the z-axis of the OCT scan; Minimum motion correlation, maximum motion delta and maximum motion factor, all of which utilise the nerve fibre layer and total retinal thickness to calculate Pearson correlation and absolute differences between the thickness values from each set of consecutive B-scans. The lowest correlation and highest absolute difference in a scan define the resulting indicator values. These values identify blinks, eye motion artefacts and segmentation failures. It should be noted that the image quality score and segmentation indicators are often correlated with one another. Finally individuals with outlier values of refractive error were removed from the study. Outlier refractive error scores were defined as values lying outside one standard deviation of 1.5 times the inter-quartile range from the median. The final dataset included 31,135 individuals.

### Genome-wide Association Studies

GWAS were implemented using an additive linear model in BGENIE [17]. The mean thickness of each component PRC layer across the ETDRS grid was used as the input phenotype (fig. 1C). Eye-specific covariates including image quality measures obtained from the OCT machine (listed above) and refractive error (calculated as *spherical error* + 0.5 × *cylindrical error*) were regressed out of thickness values for the left and right eye separately before a mean was made across the two eyes. Covariates including age, weight, sex, height, OCT machine ID and the first 20 genotype PCs were used in the model. A genome-wide significance threshold of P *<*5 × 10^−8^ was used. LD-score regression was implemented using LD SCore v1.0 [19].

#### Associated variant discovery set

The summary statistics from the GWAS of the ONL, IS and OS were meta-analysed using MTAG [20]. The resulting lowest p-value of the three meta-analysed phenotypes was considered the meta p-value. The meta-analysed PRC results were filtered using GCTA Conditional and Joint Analysis (COJO) [21] to select for independent SNPs that were more than 10Mb apart. Further filtering was applied and loci within 1.5Mb of one another were labelled as being in the same locus as indicated in the results tables where loci considered to be within the same locus are shaded the same colour, alternating grey and white (table S3). A plot of a magnified locus of interest was generated using LocusZoom [22].

### Exome analysis

We performed gene burden testing using the UK Biobank (UKBB) 200,000 whole exome release to search for genes that showed an increased level of rare loss of function (LoF) or rare missense variants that were predicated as deleterious and probably damaging. Briefly, we ran the ensemble variant effect predictor (VEP) [23] on the multi sample VCF from the 200K UKBB release to allow us to classify variants and assign their consequence. We defined high confidence LoF variants using the VEP plugin LOFTEE and selected missense variants using SIFT “deleterious” and Polyphen “probably damaging” flags, all variants were filtered to have a minor allele frequency (MAF) of less than 1% prior to collapsing into gene based genotypes for burden testing. To define gene level genotypes we used the standard collapsing method where we considered all variants within the gene for each sample separately and defined zero, one and two (0,1,2) genotypes based on: zero (genotype 0) no rare variant of the given type present in the gene; one (genotype 1) more than zero rare variants of the given type present in the gene but all are in the heterozygous state; and two (genotype 2) if any of the rare variants of the given type where in the homozygous state. It is worth noting that by using this approach we do not account for compound heterozygous mutations or the overall variant load within individual genes but it does allow us to interrogate potential rare variant associations to the trait of interest at the gene level. To perform the association testing we treated the gene level genotypes for rare LoF and potentially damaging missense variants as standard genotypes within a linear model with covariates against the OCT derived PRC layers. The covariates included within the models and the overall sample selection was exactly the same as for the common SNP GWAS analysis. A significance threshold of P *<*5 × 10^−5^ was used.

#### Annotation of variants and geneset enrichment

Significant variants were manually annotated with associated gene and any prior ophthalmic or general phenotypic associations using ENSEMBL [24] and Open Targets Genetics platform [25]. Geneset and tissue enrichment analysis was conducted using DEPICT [26].

### Comparison of Genetic Effect on Retinal Sub-fields

The foveal, intermediate and peripheral fields of the ETDRS grid were considered separately (fig. 2). A mean thickness of each of the PRC layers was calculated across each of these fields. A GWAS was then performed for each of these phenotypes. Eye specific covariates were regressed out of the individual eye measurements prior to a mean being made across left and right eyes, as described above. The same covariates were used in these GWAS as were in the simple phenotype GWAS. The GWAS were performed using BEGENIE [17].

**Fig 2.**
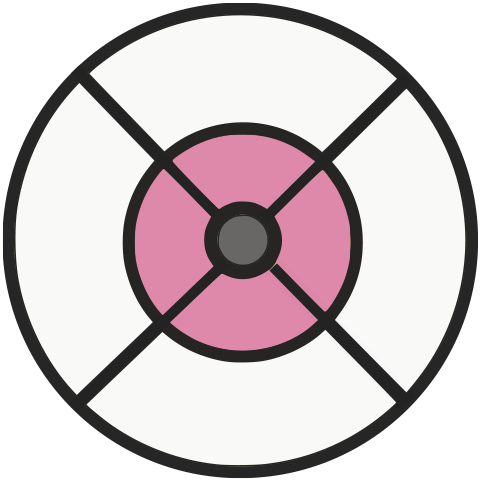
Comparison of concentric rings defined by the ETDRS grid. The macula divided into three concentric fields for comparison utilising the ETDRS grid. The peripheral field is shaded white, the intermediate field is shaded pink and the foveal field is shaded grey.

To compare the genetic effect of each SNP on the different subfields, each retinal layer was considered at a time. For each PRC layer, comparisons were made between the genetic effect on: the foveal field compared to the intermediate field; the foveal field compared to the peripheral field; and the intermediate field compared to the peripheral field. For each comparison, SNPs were selected that were significantly associated with either of the two phenotypes being considered. A z-score was calculated for each of the SNPs comparing the genetic effect size for the two phenotypes. The z-score was defined as:

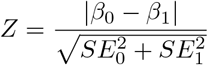

P-values were calculated for each of the z-scores and SNPs selected that were significant following bonferroni correction. The list of SNPs was clumped into loci based on linkage disequilibrium values. Each variant was annotated with all SNPs with r^2^*>*0.05 in a 1000kb window using PLINK and the locus with the lowest p-value in each window selected.

### Visualisation of spatial genetic effect

A linear model was assessed of the effect of each of the loci identified as differentially affecting the concentric retinal fields on the thickness of each of the PRC layers in each of the nine segments in the ETDRS grid. The model included the same covariates as the GWAS of average retinal thickness and the effect size was plotted back into the ETDRS grid space.

### Interaction models

Linear models were used to explore the presence of genetic interaction effecting the thickness of the three PRC layers. The same covariates were used as those used in the GWAS. Significance values were corrected for multiple testing using Benjamini and Hochberg correction and a false discovery threshold of *<* 0.15 was used.

## Results

Initially we conducted three genome wide association studies (GWAS), one for the thickness of each component PRC layer - the ONL, IS and OS - averaged across both the ETDRS grid and across both left and right eyes (fig. 1C). This is analogous to many clinical uses of macular layer thicknesses. Quality control was applied to the OCT image data consistent with previous work [9, 18] and we used a well established method to select the European-associated PCA cluster using genetic data, which aims to minimise variation in non-genetic factors and genetic factors (See Methods). The resulting subset of samples that passed both image and genetic filters comprised of 31,135 individuals (table S1). The three GWAS (one for each component layer of the PRC), each identified a number of associated loci using the established genome-wide significance level of P *<*5 × 10^−8^ threshold. Following calculation of the linkage disequilibrium score for each of the three GWAS, there was minimal evidence for inflation due to residual population structure (fig. S1, table S2). SNP heritability of the thickness of each of the PRC layers was calculated: ONL h^2^=0.39, IS h^2^=0.18, and OS h^2^=0.21. Unsurprisingly we observed numerous loci that were significantly associated with the thickness of more than one of the PRC layers, and so we meta-analysed results using MTAG [20]. Following selection of independent loci using COJO [21], there were 111 loci significantly associated with the meta-analysed PRC layers’ thickness (Figure fig. 3, table S3 and table S4).

**Fig 3.**
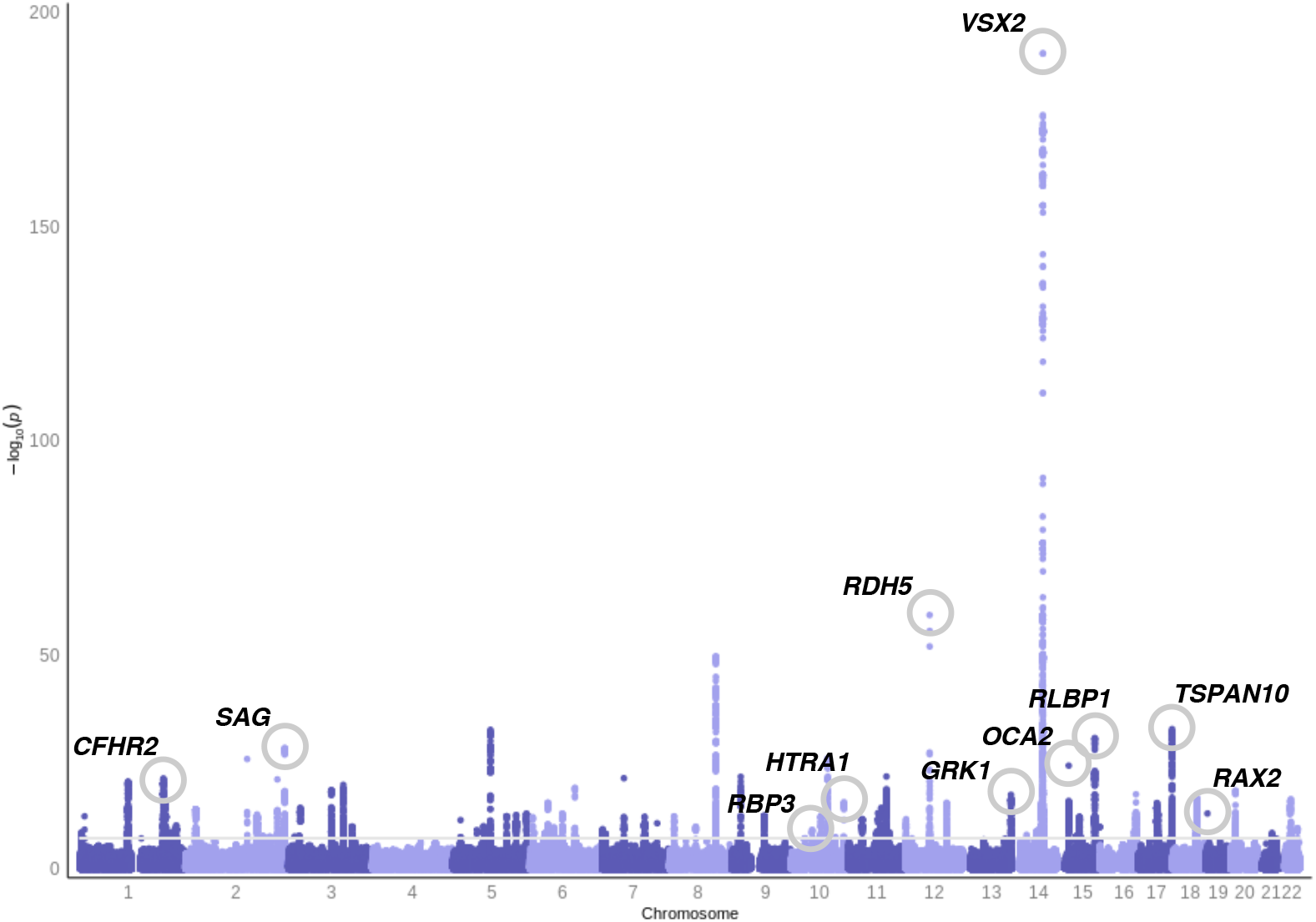
Genome-wide association study of photoreceptor cell layer thickness phenotypes. Manhattan plot of the photoreceptor cell layer thickness phenotype GWAS p-values. These result from meta-analysis using MTAG [20] of the summary statistics from GWAS of the thickness of the outer nuclear layer (ONL), inner segment (IS) and outer segment (OS). Variants are considered significantly associated if they reach genome wide significance (P *<*5 × 10^−8^). Loci of interest, selected through manual labelling by ophthalmic experts, are circled and labelled.

We immediately noticed that many of these loci were close to well known genes associated with rare ocular diseases (fig. 4 and table S5). 17 of the 111 loci overlap with a known rare retinal disease locus, which is highly unlikely to occur by chance (P *<*2.2 × 10^−16^ Hypergeometric distribution).

**Fig 4.**
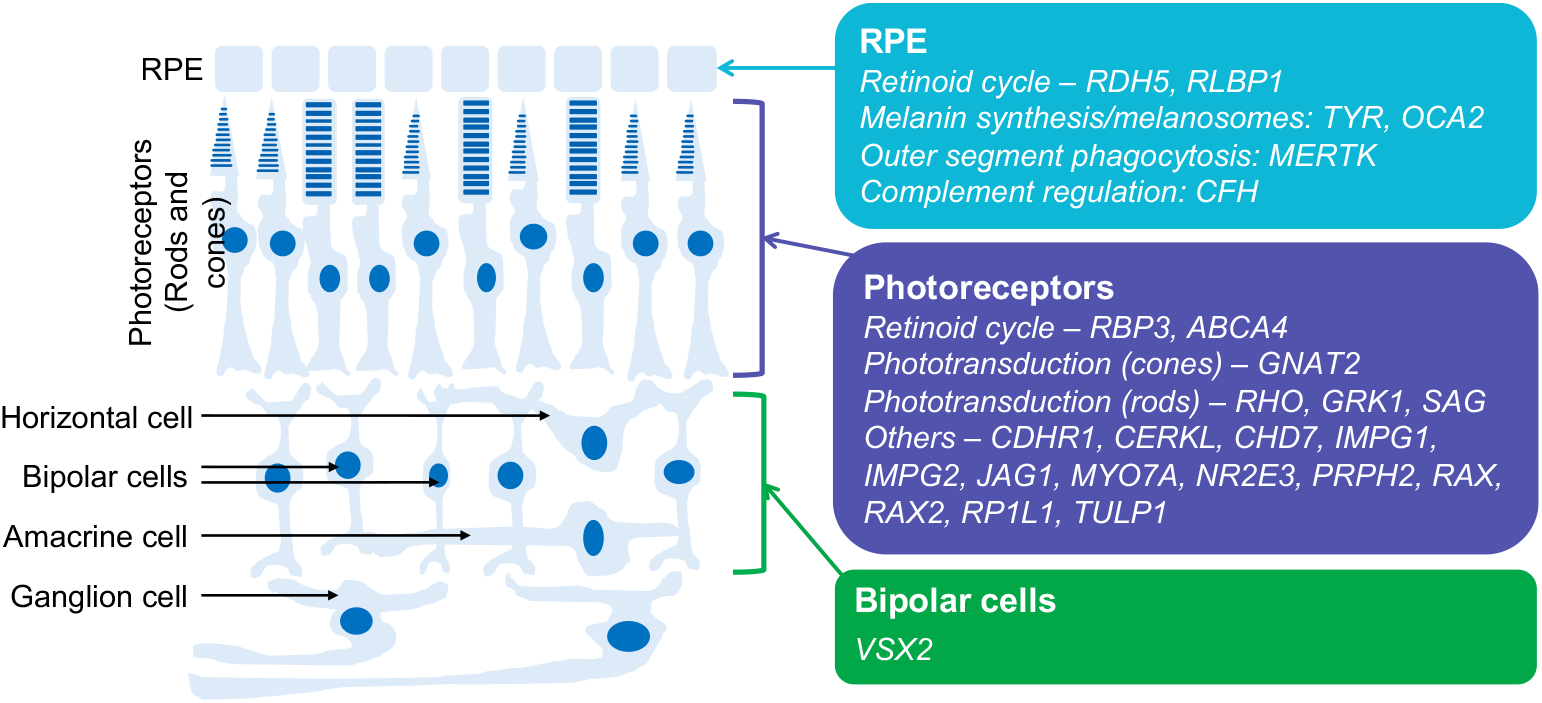
Schematic of retina showing main cell types with sites of expression (in mature human retina) of many of the genes associated with rare monogenic ocular disease found during genetic discovery (table S5). Some genes are also expressed significantly in other cell types not shown (e.g. *RLBP1* is expressed also in Muller cells). Also, some genes (including those not shown) have a wider role in development, but expression is more restricted in adults (e.g. *VSX2* is expressed in retinal progenitor cells in development, but mainly in bipolar cells in the mature retina).

We further leveraged the initial release of the UK Biobank exome dataset of 247,000 individuals, complementing the common variation GWAS with a rare variation gene burden test on the same phenotypes. We considered both strict loss of function mutations and overall missense mutations as two separate tests applied to each layer. Although there were no strong rare variant hits, setting a more liberal significance threshold (P *<*5 × 10^−5^) - consistent with other exome wide burden tests - we found 10 significant loci (table S6,fig. S2). Three of these loci overlap with rare eye disease, again unlikely to have been observed by chance (P=2.65 × 10^−6^ by hypergeometric distribution).

Overall between rare and common discovery we have found over 100 loci associated with PRC thickness in some manner, and these continuous quantitative phenotype associations have significant overlap to known rare ocular disease genes.

### Selected Loci of Interest

Of these genetic variants, the locus with the smallest p-value and with a reasonably large effect size for a single locus was *VSX2. VSX2* is a homeobox transcription factor that has been found expressed in the developing retina of humans, mice and zebrafish [27]. The gene is known to have an important role in ocular size, with pathogenic variants leading to microphthalmia [27]. This locus showed allelic heterogeneity in the GWAS results with the most significant genetic variant being rs1972565 (P = 5.23 × 10^−191^) whilst COJO analysis retained 4 other SNPs across the *VSX2* locus. In addition, there are SNPs in nearby genes which could be long range regulatory SNPs for *VSX2* rather than the closest genes we have assigned this to (fig. 5). Interestingly in previous work we identified genetic variants in neighbouring loci that had a statistically significant effect on the thickness of the inner retinal layers [9]. All this evidence points towards the *VSX2* locus having multiple alleles which affect different aspects of eye development.

**Fig 5.**
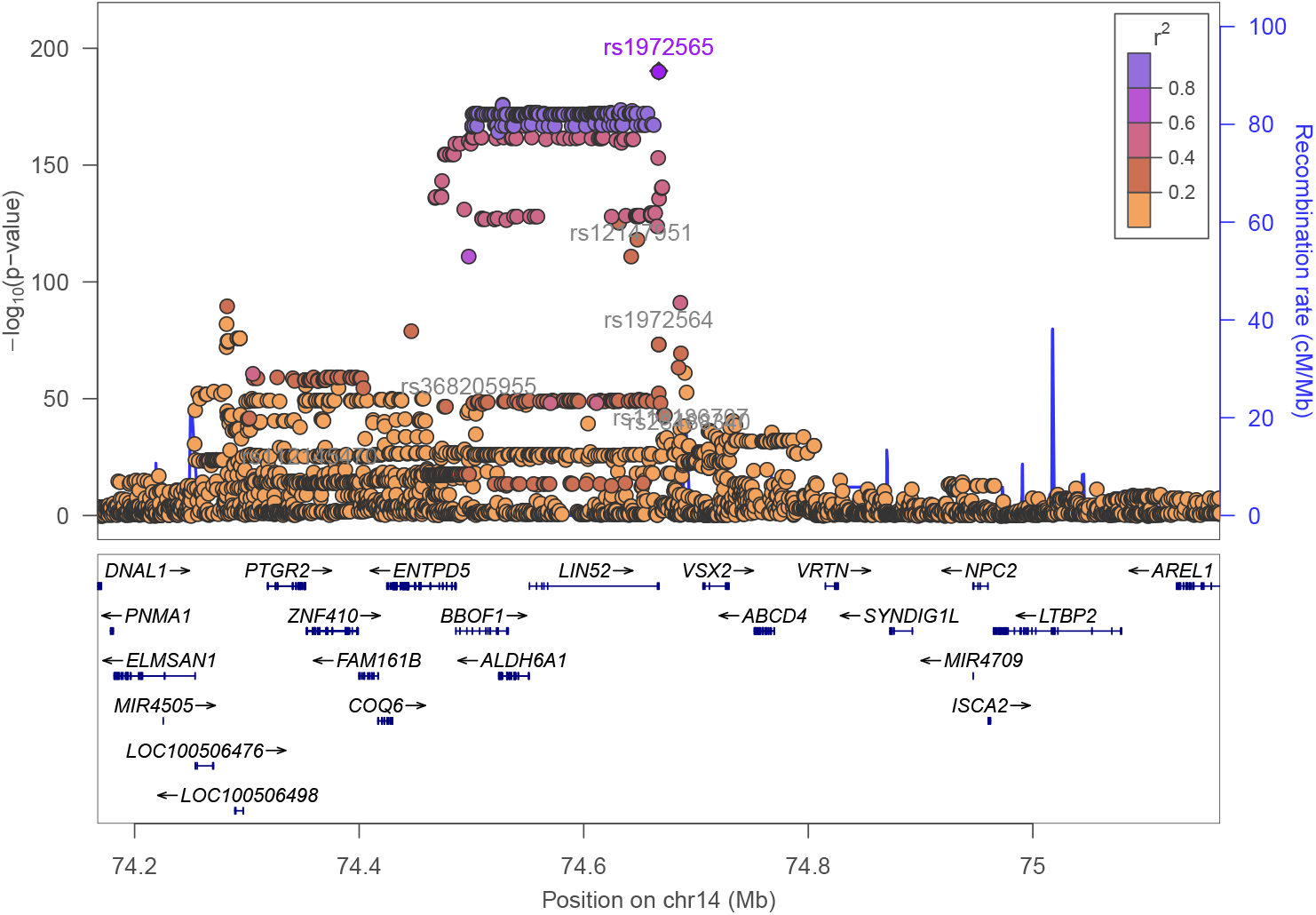
Magnification of the *VSX2* locus. Association of the locus on chromosome 14 centred on the genetic variant rs1972565 (highlighted in purple). Other genetic variants found significantly associated with PRC thickness are labelled in grey.

Further there were several loci found associated with PRC layer thickness that are known to be involved in the retinoid cycle. The retinoid or visual cycle refers to the recycling of chromophore: photons of light isomerise 11-*cis* retinal to all-*trans* retinal in the photoreceptor outer segments; to be converted back to 11-*cis* retinal, a multi-stage process is entailed, which includes transport to the retinal pigment epithelium and then back to the photoreceptor [28]. *RDH5, RLBP1* and *RBP3*, all found associated with PRC layer thickness, each have a known role in the retinoid cycle and are associated with rare inherited retinal diseases. *RDH5* (rs3138142, P = 5.58 × 10 ^-60^) encodes 11-*cis* retinol dehydrogenase, an enzyme responsible for conversion of 11-*cis* retinol to 11-*cis* retinal. The specific SNP in *RDH5* (rs3138142, P = 5.58 × 10 ^-60^) has previous associations with a number of ophthalmic phenotypes including the age one started wearing glasses, cataract and myopia. Pathogenic variants in this gene are known to cause fundus albipunctatus, a retinal dystrophy and form of night blindness that primarily affects the rods [29, 30]. *RLBP1* (rs3825991, P = 2.70 × 10^−31^) encodes the cellular retinaldehyde binding protein, a transporter of 11-*cis* retinal [31, 32]. Mutations in this gene lead to a number of different retinal dystrophies [33]. *RBP3* (rs111245635, P = 8.41 × 10^−10^) encodes the interphotoreceptor retinoid-binding protein which has a role in transporting retinoids between the RPE and the photoreceptor cells [34]. Additionally, rare variants within *RPB3* cause recessive retinal dystrophy and high myopia [35]. Much of the retinoid cycle occurs in the supporting RPE layer, for example *RDH5* is almost exclusively expressed in this layer [36], showing that one does not need gene expression in a cell type to impact its structure.

Two loci, *SAG* (rs7564805, P = 6.16 × 10^−29^) and *GRK1* (rs9796234, P = 4.69 × 10^−18^), were found associated with PRC layer thickness and these genes have known involvement in Oguchi disease. Oguchi disease is a rare autosomal recessive disease. Features include the Mizuo-Nakamura phenomenon, a golden retinal sheen and night-blindness [37]. *SAG* encodes arrestin and *GRK1* encodes rhodopsin kinase; both proteins are involved in shut-off of activated rhodopsin, enabling termination of the light response. When these proteins are dysfunctional, rhodopsin continues to activate the phototransduction cascade, shutting off the cyclic nucleotide-gated current, hyperpolarising the rods, and rendering them electrically unresponsive to further light photons. Thus patients primarily experience night blindness in this disorder. Studies have also found changes in the microstructure of the retinal layers in Oguchi disease patients [38, 39]. The presence of common variation in these genes suggests that there might be functional differences in rod physiological due to genetics present at the population level.

Further, several variants were found in the large heterogenous *NPLOC4-TSPAN10* locus, and both rare and common variants were discovered in the *OCA2* locus. Variants in these loci have previously been found associated with foveal structure [9] suggesting that overall retinal development has an impact on the PRC layer. Additionally the *NPLOC4-TSPAN10* locus is associated with strabismus, the abnormal alignment of the eyes [40]. Strabismus is known to be caused by defects in the extraocular muscles, cranial nerves, refractive error and visual cortex processing. Our finding suggests there may be a role of retinal structure in strabismus which warrants further investigation utilising appropriate clinical datasets.

We also note that several loci significantly associated with the PRC layer thicknesses had prior associations to AMD. These included *CFHR2* (rs410895, P=8.19 × 10^22^), *HTRA1* (rs60401382, P=2.43 × 10^−16^) and *RAX2* (rs76076446, P=1.07 × 10^−13^). AMD is a disease characterised by the atrophy of PRCs caused by the accumulation of lipid deposits in the RPE. Extensive recent work has previously been completed to study the relationship between PRC thickness and AMD [41, 42].

The discovery of loci associated with PRC thickness at genes encoding proteins involved in phototransduction, generating the photoreceptor electrical response to light, is of interest. These include *GNAT2, SAG, GRK1* and *RHO*. This suggests there is a shared importance of these genes between phototransduction and structure. It may warrant further investigation into the relationship between the structure and function of these cells and these variants might provide interesting genetic instruments for the integration of the physiological process of light transduction with other features.

To further explore the biological pathways underlying PRC morphology, we used DEPICT [26] to perform gene set and tissue enrichment analysis. When applied to the loci associated with PRC thickness, numerous genesets reached statistical significance following false discovery rate (FDR) correction (table S7). These include ocular-associated genesets such as *abnormal lens morphology, microphthalmia, disorganized retinal layers* and *abnormal ocular fundus morphology* amongst others. Additionally, tissue enrichment analysis of the loci associated with PRC thickness resulted in significant associations with *retina* and *eye* tissue (table S8).

Additionally, there were a number of genes identified in the gene-burden test analysis with interesting prior associations. These include *OCA2*, also significantly associated in our GWAS analysis, which has a prior association to oculocutaneous albinism. Individuals with this condition are characterised by foveal hypoplasia, the absence of the formation of the foveal pit. Further, three genes, *ABCA4, MYO7A* and *NR2E3*, are shown to have an association to PRC thickness and have a prior association with retinal dystrophies. *ABCA4* encodes ATP-binding cassette sub-family A member 4, a protein expressed almost exclusively in the PRCs. It is responsible for mediation of all-trans-retinal aldehyde across the PRC membrane, clearing it from the retina following phototransduction. ABCA4 is the gene most frequently implicated in monogenic retinal dystrophies [5, 43]. Biallelic pathogenic variants are the most common cause of of juvenile macular degeneration (Stargardt disease) and can also give rise to a cone-rod dystrophy. *MYO7A* encodes myosin VIIA, a protein found in inner ear and retina. Within the PRCs, this protein helps regulate transport of opsins, light-sensitive proteins required for vision. Mutations in *MYO7A* are known to cause Type 1 Usher Syndrome, a condition characterised by deafness and retinal degeneration [44]. *NR2E3* encodes a nuclear receptor transcription factor that activates rod development and represses cone development [45]. Mutations in this gene are also associated with retinitis pigmentosa in addition to enhanced S-cone syndrome [46].

### Fine grained topography of genetic variation

In addition to the clinically utilised measure of the mean PRC thickness taken across the ETDRS grid, TABS outputs the thickness of each of the PRC layers within each of the nine ETDRS subsections. To capitalise on this increased dimensionality of data, we explored how genetic variation affects PRC morphology in the different areas of the ETDRS segmentation grid. We chose to consider the macular field split into three concentric fields which align with the topography of the valley-like macula (fig. 2). A mean thickness across each of these concentric fields - the foveal, intermediate and peripheral fields - was taken for each of the PRC layers. These phenotypes were used as an input to GWAS and the SNPs which passed the significance threshold of 5 × 10^−8^ were taken forward for further analysis. Z-score analysis was applied to the resulting effect size to identify loci with significantly different effect on thickness of the different concentric fields of each PRC layer. Numerous (32) loci were found in each layer that affect the thickness of the concentric fields differentially (fig. 6, fig. S3).

**Fig 6.**
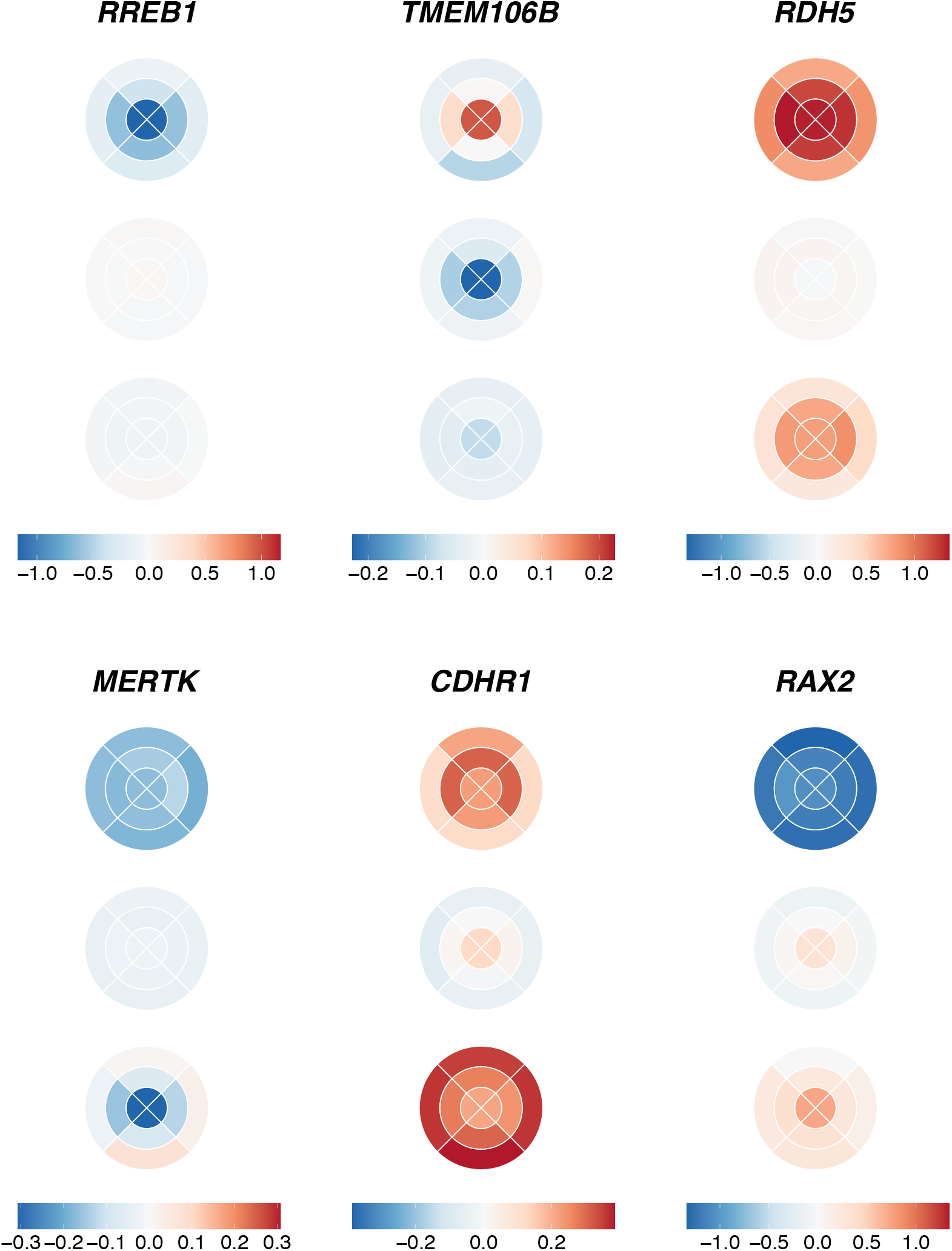
Effect size of selected SNPs on ETDRS grid. The figure shows the effect size of the SNPs (shown by a blue-red gradient) with differential effect on the concentric fields of the retina, on the nine different segments of the ETDRS grid across the three PRC layers listed from top to bottom: ONL, IS and OS. Six loci are visualised: *RREB1* (rs75757892), *TMEM106B* (rs13237518), *RDH5* (rs3138142), *MERTK* (rs869016), *CDHR1* (rs55798570), and *RAX2* (rs76076446). Effect sizes are scaled for each genetic locus individually.

The locus showing the most significant difference in effect on the thickness of the ONL across the concentric fields was *RREB1* (rs75757892) which had a differential effect on the foveal field (effect size=-1.12) compared to both the intermediate field (effect size=-0.55) and peripheral field (effect size=-0.20). *RREB1* encodes a zinc finger transcription factor, the Ras-responsive element binding protein 1 [47]. Genetic variants at *RREB1* have prior associations to various ocular phenotypes including the age one starts wearing glasses and spherical power.

The locus showing the most significantly different genetic effect on the IS thickness across the macula was *TMEM106B* (rs13237518) which had a significant difference between the foveal field (effect size=-0.23), and the peripheral field (effect size=-0.02). *TMEM106B* is known to be involved in frontotemporal lobar degeneration, a neurodegenerative disease [48]. There is growing interest in the use of OCT derived retinal thickness measures as biomarkers for neurodegenerative diseases [49, 50] making this finding of potential interest.

The locus with the most significantly different genetic effect on the thickness of the OS was *RDH5* also found in the initial meta-GWAS. *RDH5* (rs3138142) shows a significantly different effect on the thickness in the peripheral field (effect size=0.81) compared to both the foveal field (effect size=1.37) and the intermediate field (effect size=1.28). As described above *RDH5* encodes 11-*cis* retinol dehydrogenase, an enzyme that is part of the retinoid cycle. It has been previously associated with the retinal dystrophy, fundus albipunctatus.

Notably several further loci were discovered to differentially affect the concentric fields of the retina that had prior associations with retinal dystrophies. These include *MERTK* (rs869016) which shows a significant difference of effect across the concentric fields of the OS. It has a significantly larger effect size on the fovea (effect size=-0.31) as compared to the peripheral field (effect size=0.02). *MERTK* encodes MER proto-oncogene, tyrosine kinase, a transmembrane protein localised to the RPE. *MERTK* has a known role in the phagocytosis of the OS by the RPE. Mutations in *MERTK* have been associated with retinitis pigmentosa and early onset retinal dystrophies [51].

Additionally *CDHR1* (rs55798570) shows a significantly different effect size on the thickness of the IS in the foveal field (effect size=0.13) compared to both the intermediate field (effect size=0.002) and the peripheral field (effect size=-0.06). *CDHR1* encodes cadherin-related family member 1, a cell adhesion protein that localises to the junction of the IS and OS. Genetic variants in *CDHR1* have been associated with a number of inherited retinal dystrophies including retinitis pigmentosa and cone-rod dystrophy [52].

Further *RAX2* (rs76076446), shows a different effect on the thickness of the IS at the fovea (effect size=0.31) compared to the peripheral field (effect size=-0.09). *RAX2* encodes retina and anterior neural fold homeobox 2, which has a known role in eye development. Pathological variants in this locus are known to cause cone and rod dysfunction and retinitis pigmentosa, a rod-cone disease [53].

Each of these retinal dystrophies have distinct and sometimes opposing spatio-temporal patterns in which they affect the rods and cones. Some dystrophies, including retinitis pigmentosa, affect the rods first, with the cones affected later. Patients have problems with night vision, then peripheral vision, and finally central vision. Fundus albipunctatus also affects rod function, whilst cones are usually spared. Others such as cone-rod dystrophy affect the cones first and the rods afterwards. Cones are found at the highest density at the fovea, whilst rod density peaks more peripherally. There appears to be a parallel between the spatial genetic effects of these loci on thickness of the PRC and the organisation of PRCs affected by the diseases associated with these loci.

### Genetic Interactions

As there are both clear common biological pathways associated with multiple discovered variants, and overlaps with known rare disease loci we wanted to explore whether genetic interactions were affecting the retinal phenotype. Genetic interactions of common variation are challenging to discover and characterise due to the large space of possible interactions. To restrict our space we considered interactions in the following categories: (1) Interactions of the developmentally important VSX2 locus internally and with all other common genetic variants; (2) Interactions between the Retinoid cycle loci; Interactions of the Oguchi syndrome loci; (4) Interaction of all loci with genetically defined sex; and (5) Interactions between rare burden levels in genes and nearby common SNPs.

Overall we do not find widespread interaction terms that would pass multiple testing criteria. There was no statistically significant interaction term between the genetic variants with prior association to Oguchi disease, or between those with a prior association with the retinoid cycle. Additionally, there was no evidence of an interaction between any of the 111 identified loci and genetically defined sex. There was no evidence for interactions between rare burden levels in genes and nearby common SNPs; however this analysis was limited by the sample size of individuals with heterozygous alternative allelic states. Expansion of this analysis in a cohort containing a higher proportion of individuals with rare disease phenotypes would have more power to identify interactions.

However, following Benjamini and Hochberg correction for multiple testing, and using a False Discovery threshold level of *<* 0.15, four variants show an interaction with the lead *VSX2* SNP, rs1972565 in relation to their effect on ONL thickness. Two of the four variants (rs112145470 and rs28488340) are within the same locus as *VSX2*, consistent with the already noted allelic hetreogenity at this locus. The other two loci are *DYNLRB2* (rs7206532) and *PRPH2* (rs375435). *DYNLRB2* is expressed in retinal Muller cells and astrocytes, and encodes dyenin light chain roadblock type 2. Mutations in this gene are associated with deafness and hepatocellular cancer. *PRPH2* encodes peripherin 2 which is localised to the photoreceptors. Mutations in this gene are known to result in forms of retinal dystrophy and macular degeneration [54]. Upon stratification of the population by their genotype at *VSX2* and *PRPH2*, a change in ONL thickness can be observed, specifically between *PRPH2* homozygous reference and homozygous alternative genotype in those with the *VSX2* homozygous reference genotype (fig. 7,fig. S4). In individuals with *PRPH2* homozygous reference, a thinner ONL is found in the superior segments of the macula, and thicker ONL in the inferior segments. In individuals with *PRPH2* homozygous alternative, the inverse pattern is seen. This is of particular interest as there is often a difference in PRC degeneration across the macula eqautor in retinal dystrophies. *PRPH2* has a doccumented specturm of penetrance and associated disease phenotypes [55, 56]. This finding suggests one of the possible mechanisms by which the effects of variants at the *PRPH2* locus are modified is via the genotype at the *VSX2* locus.

**Fig 7.**
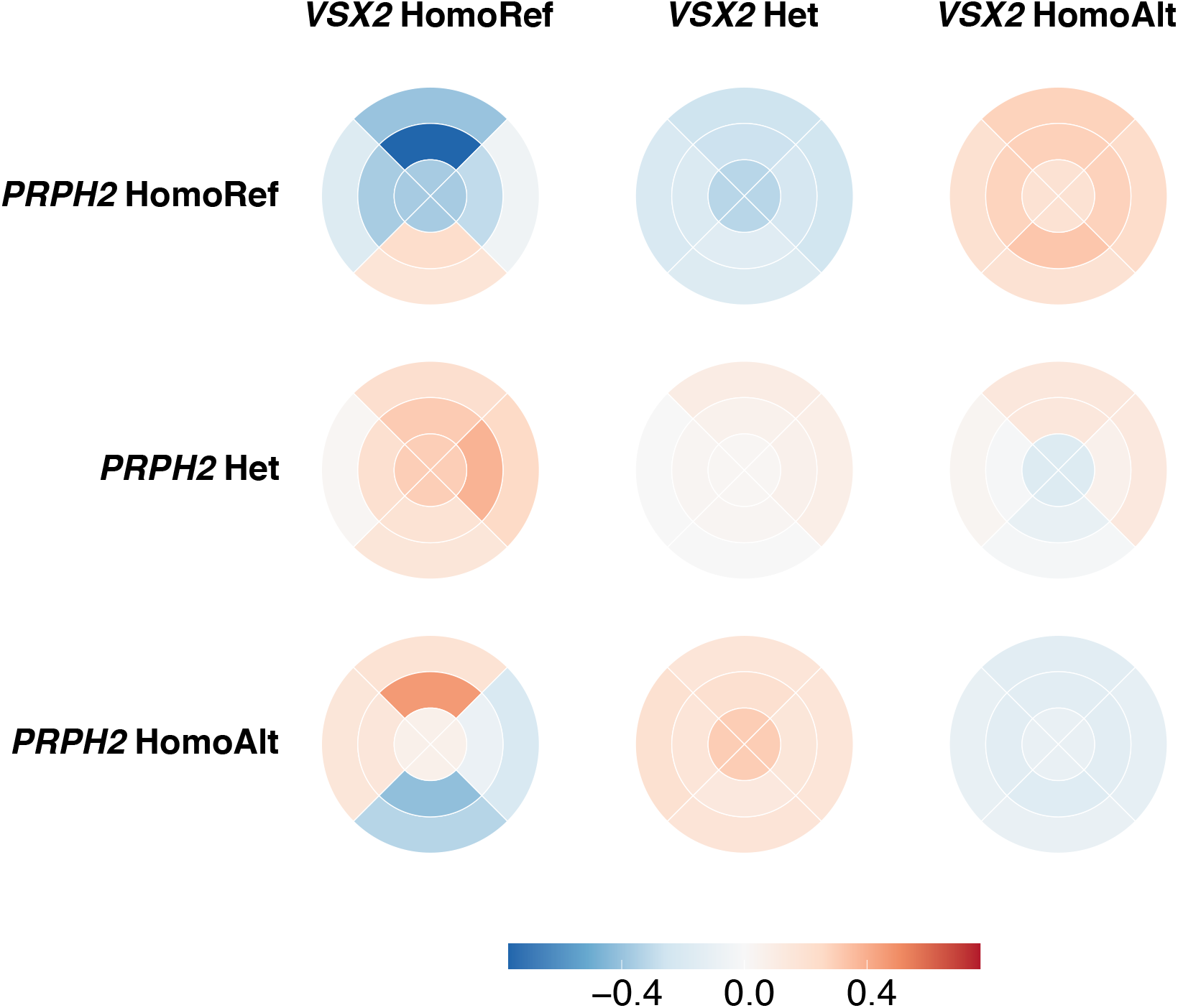
Normalised thickness of ONL stratified by *VSX2* and *PRPH2* genotype. The mean thickness of the ONL at each of the nine segments of the ETDRS grid across individuals stratified by their genotype at *VSX2* and *PRPH2*. The thickness is mean normalised per *VSX2* genotype.

## Discussion

Here we have performed the largest and most detailed GWAS and rare variation association of PRC layer thickness to date, leveraging the high-dimensional spatial resolution available within retinal OCT. We initially explored a simple phenotype of the mean thickness of each of the PRC layers - the ONL, IS, OS - across the entire macula field as defined by the ETDRS grid. These phenotypes are advantageous in that they are aligned with those used in clinical practice. Using these phenotypes, we identified 111 independent loci significantly associated with one or more of the component PRC layers following GWAS meta analysis and 10 loci associated with rare variation following gene burden testing. Many of the loci identified here had prior associations to ocular phenotypes and the loci collectively were enriched for known rare ocular diseases.

The collection of rare disease loci discovered in this study of common, physiologically healthy individuals without obvious eye diseases shows the continuum between common and rare variation, and rare variation which leads to disease. In this study we explicitly looked for interactions between common variation at different loci and common and rare variation at the same loci. We found several relatively weak interactions between a SNP near the well known eye development gene, *VSX2*, and two SNPs within this locus as well as two SNPs near *PRPH2* and *DYNLRB2*. No evidence was found for an interaction between common and rare genetic variants but this was partially due to limitations of the dataset and its absence of adequate numbers of individuals with rare variation and subsequent lack of statistical power.

Genetic discovery also found sets of loci associated with PRC thickness with prior associations to different ocular phenotypes. Four loci (*GNAT2, SAG, GRK1* and *RHO*) have a prior association to phototransduction, the generation of an electrical response to light in the PRC. Further three variants were found in or near genes (*RDH5, RLBP1* and *RBP3*) with prior associations to the retinoid cycle, the process by which the chromophore is regenerated following photoisomerisation by light. In both cases, this finding suggests a relationship between the form and function of these cells which may warrant further investigation. Whilst it might be intuitive that genes encoding structural and matrix proteins are associated with photoreceptor layer thicknesses, it is interesting that genes encoding proteins involved in bringing about or shaping the photoreceptor electrical light response to light are also implicated, potentially suggesting a role for visual signalling in development or maintenance of outer retinal structure.

There were several loci associated with specific disease. For example both *SAG* and *GRK1* in addition to an association with PRC thickness have a known involvement in Oguchi disease, a rare autosomal eye condition which causes visual impairment. As well as the overlap between rare eye genetic disease and these common variants affecting retinal structure, there are some interesting, less expected findings. For example, three loci - *MYEOV* (rs10737153), *TNS1* (rs201030469) and *FGFR2* (rs17102399) – have impacts on both PRC layer thickness and risk of diagnosis for breast cancer. Additionally *TMEM106B*, which had a particularly notable spatial genetic effect, has a known association to a form of dementia. In both cases, the link between morphology and disease suggests the possibility of using OCT derived PRC morphology measures as biomarkers for complex diseases in the future. Additional datasets enriched for individuals with these diseases with OCT data will be required to further explore these concepts.

This study has offered the ability to study the genetics underlying complex retinal structures. Whilst OCT does not directly offer single-cell resolution, the quantitative values extracted from the images in the photoreceptor layer describe a single cell layer and capture variation in sub-cellular structures. It is interesting to note that some of the strong hits have gene expression in a different cell type. For example *RDH5* which is strongly associated with thickness of all three PRC layers is expressed in the RPE [57]. Therefore we are identifying genetic variation impacting sub-cellular morphology in a between cell type manner, highlighting the complex route between genetic variation, gene function and ultimate physiological differences. Imaging offers a methodology for interrogating such processes at a high-dimensional scale in a non-invasive manner and the protocol used here could be applied to other organs of the human body.

## Data Availability

The genetic and phenotypic UK Biobank data are available upon application to the UK Biobank.

## Data Availability

The genetic and phenotypic UK Biobank data are available upon application to the UK Biobank.

## Acknowledgments

The authors are extremely grateful for the selfless participation of individuals in the UK Biobank used in this study and the staff managing these cohorts. HC has received support from EMBL and is supported by Novo Nordisk Foundation (grants NNF17OC0027594 and NNF14CC0001). APK is supported by a UKRI Future Leaders Fellowship and an Alcon Young Investigator Award. TWF and EB are supported by EMBL. OAM is supported by the Wellcome Trust (206619/Z/17/Z). The UK Biobank Eye and Vision Consortium members are: Naomi Allen, Tariq Aslam, Denize Atan, Sarah Barman, Jenny Barrett, Paul Bishop, Graeme Black, Tasanee Braithwaite, Roxana Carare, Usha Chakravarthy, Michelle Chan, Sharon Chua, Alexander Day, Parul Desai, Bal Dhillon, Andrew Dick, Alexander Doney, Cathy Egan, Sarah Ennis, Paul Foster, Marcus Fruttiger, John Gallacher, David Garway-Heath, Jane Gibson, Jeremy Guggenheim, Chris Hammond, Alison Hardcastle, Simon Harding, Ruth Hogg, Pirro Hysi, Pearse Keane, Peng Tee Khaw, Anthony Khawaja, Gerassimos Lascaratos, Thomas Littlejohns, Andrew Lotery, Phil Luthert, Tom MacGillivray, Sarah Mackie, Bernadette McGuinness, Gareth McKay, Martin McKibbin, Tony Moore, James Morgan, Richard Oram, Eoin O’Sullivan, Chris Owen, Praveen Patel, Euan Paterson, Tunde Peto, Axel Petzold, Nikolas Pontikos, Jugnoo Rahi, Alicja Rudnicka, Naveed Sattar, Jay Self, Panagiotis Sergouniotis, Sobha Sivaprasad, David Steel, Irene Stratton, Nicholas Strouthidis, Cathie Sudlow, Zihan Sun, Robyn Tapp, Dhanes Thomas, Emanuele Trucco, Adnan Tufail, Ananth Viswanathan, Veronique Vitart, Mike Weedon, Katie Williams, Cathy Williams, Jayne Woodside, Max Yates, Jennifer Yip, Yalin Zheng.

## Author Contribution

HC conceived the analysis, performed the analysis and wrote the paper. TWF performed analysis and wrote the paper. PJP, APK and ARW provided advice on analysis. OAM performed analysis and oversaw the work. EB conceived the analysis, wrote the manuscript and oversaw the work. All authors reviewed the final manuscript.

## Supporting information

**Table S1.** Comparison of biological characteristics between whole UK Biobank population with OCT data (n = 67,321), the group that passes our quality control criteria (n = 31,135), and the group that fails (n = 36,186). Results are presented as mean ± standard deviation.

|  | Total OCT Population | Pass filter | Fail filter |
| --- | --- | --- | --- |
| Height (cm) | 168.68 ± 9.25 | 169.43 ± 9.15 | 168.03 ± 9.29 |
| Age (years) | 57 ± 8 | 57 ± 8 | 57 ± 8 |
| Weight (Kg) | 78.12 ± 16.02 | 78.47 ± 15.82 | 77.82 ± 16.17 |
| Sex (m/f) | 29,713/34,929 | 14,696/16,439 | 15,017/18,490 |
| Refractive Error Left (Dioptres) | -0.32 ± 2.73 | 0.23 ± 1.49 | -0.80 ± 3.41 |
| Refractive Error Right (Dioptres) | -0.38 ± 2.73 | 0.18 ± 1.48 | -0.87 ± 3.40 |

**Table S2.**
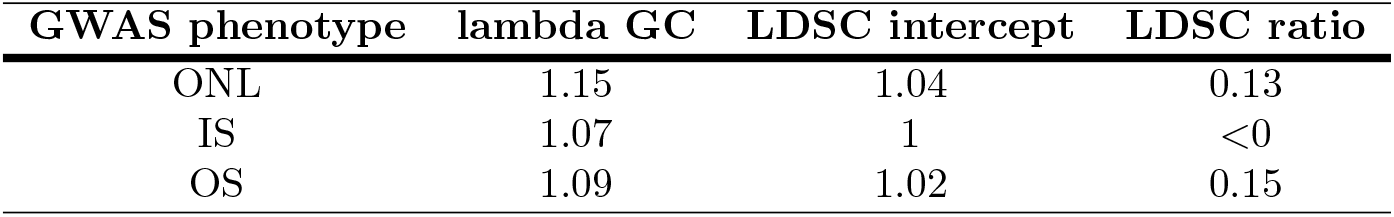
Linkage score disequilibrium analysis results. Linkage score disequilibrium metrics for the three GWAS of the photoreceptor layers following analysis using LDSCore [19]. Metrics include lambda genomic control (GC), linkage disequilibrium score (LDSC) intercept and LDSC ratio.

**Table S3.**
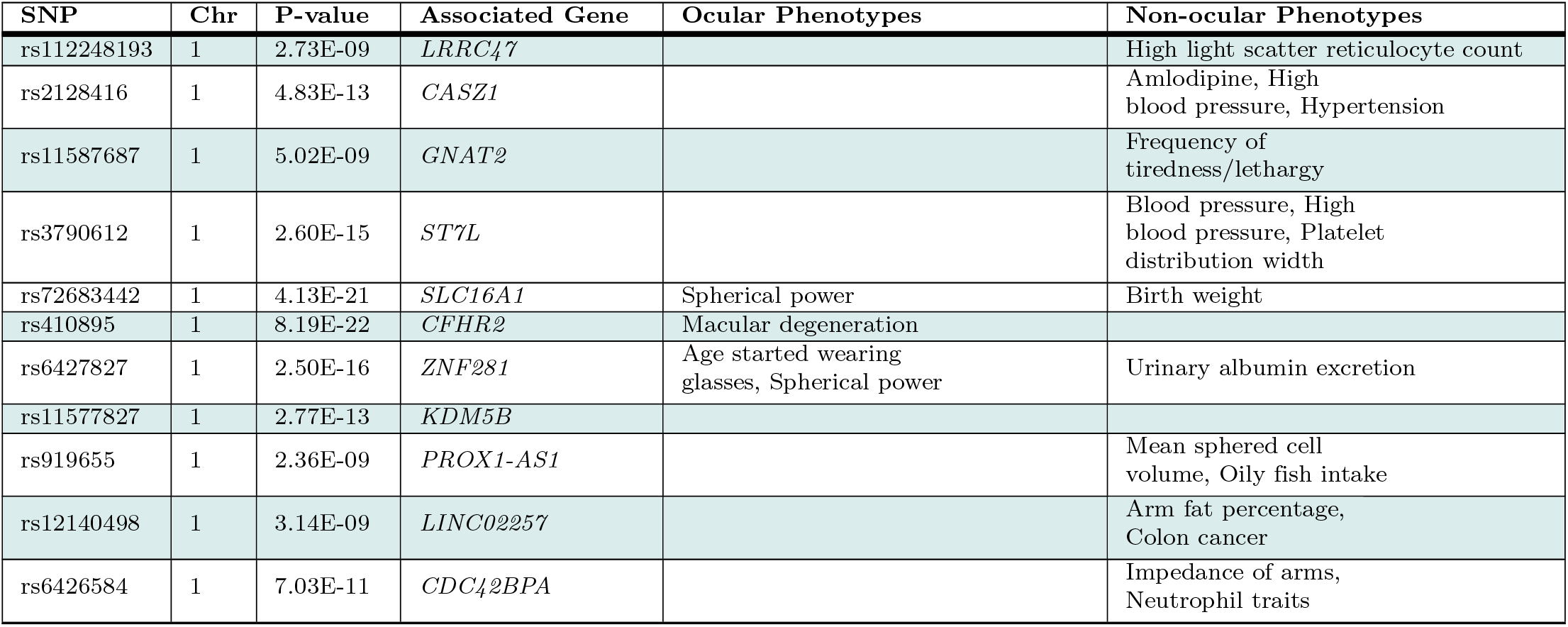
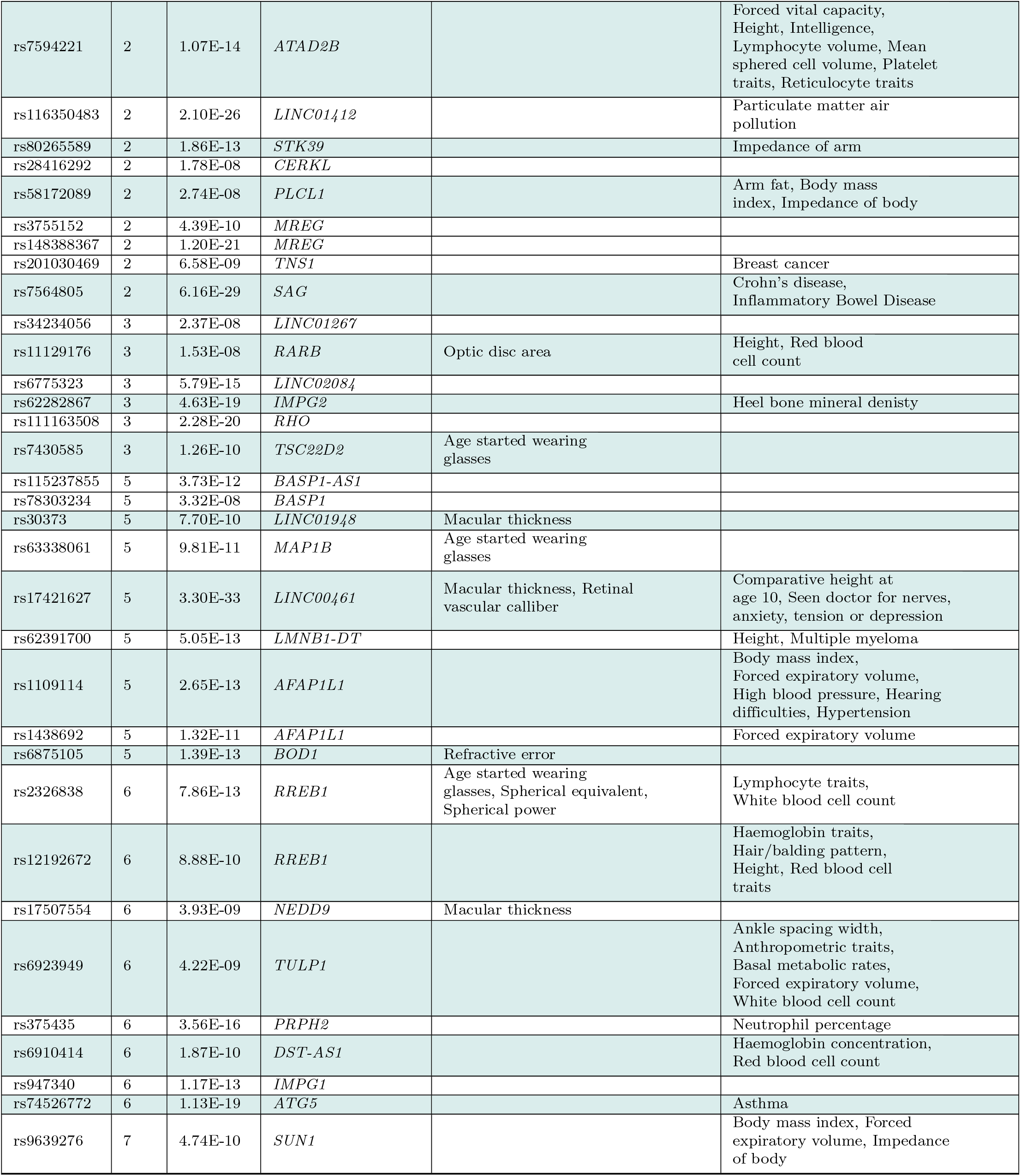
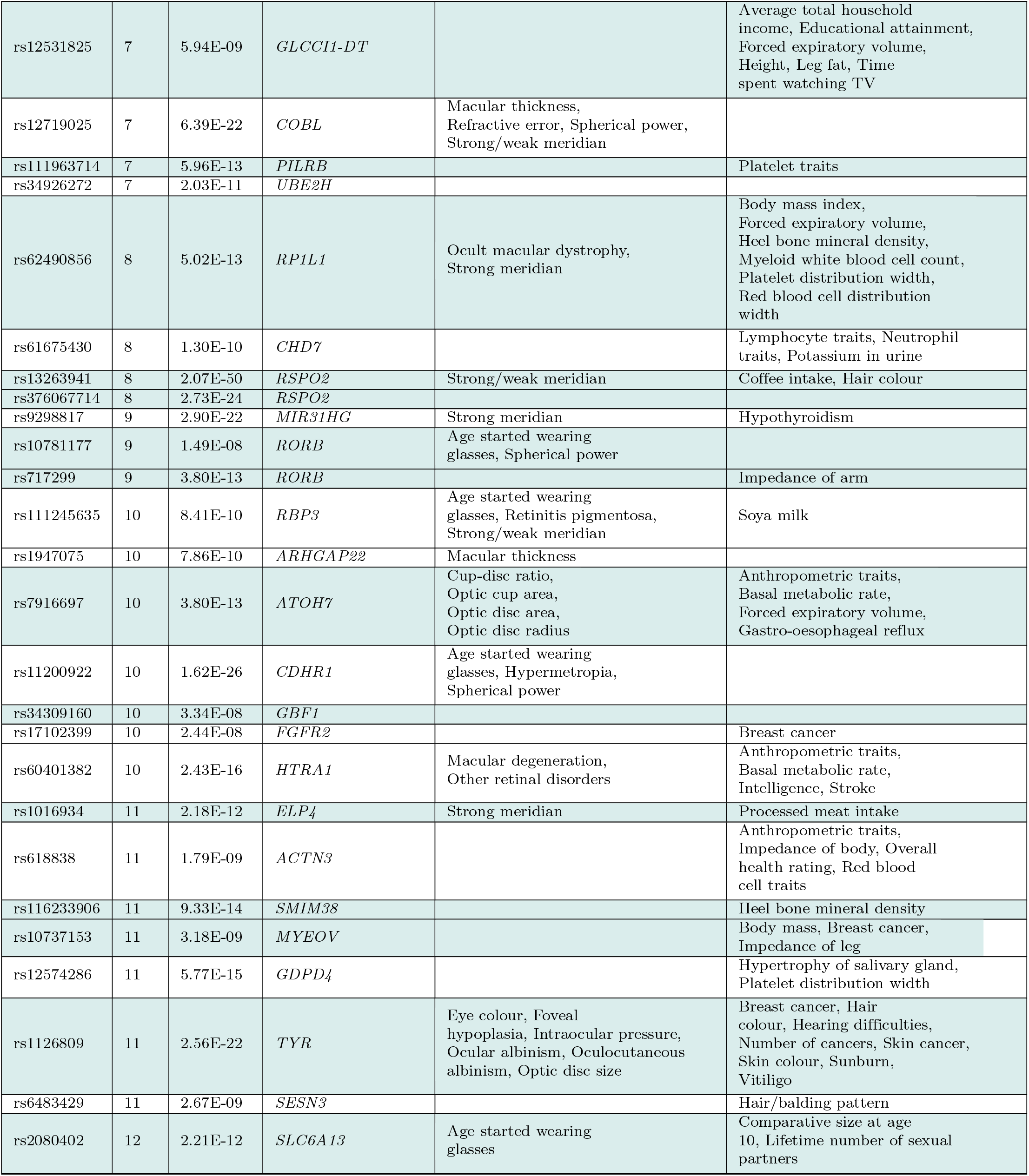
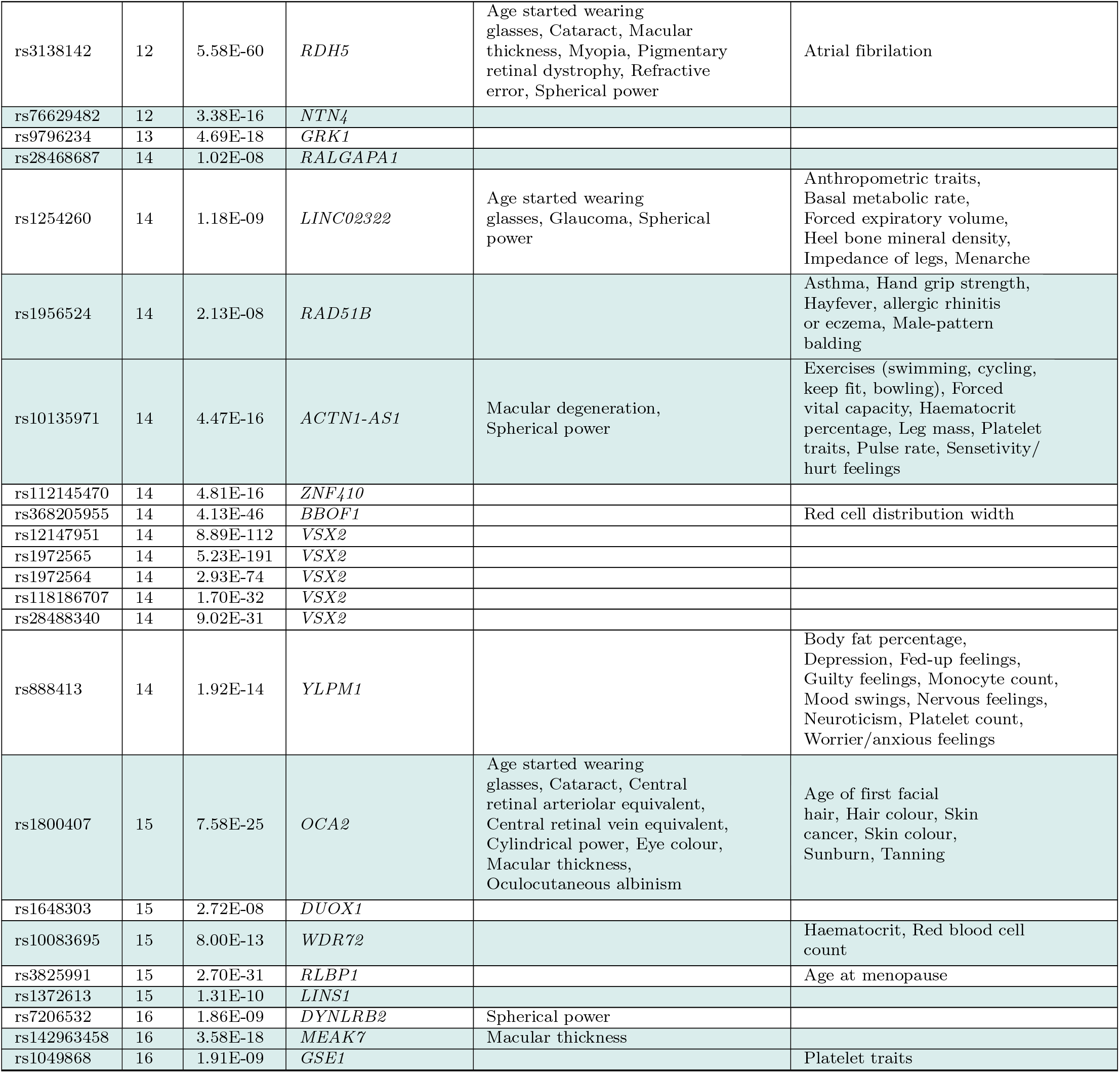
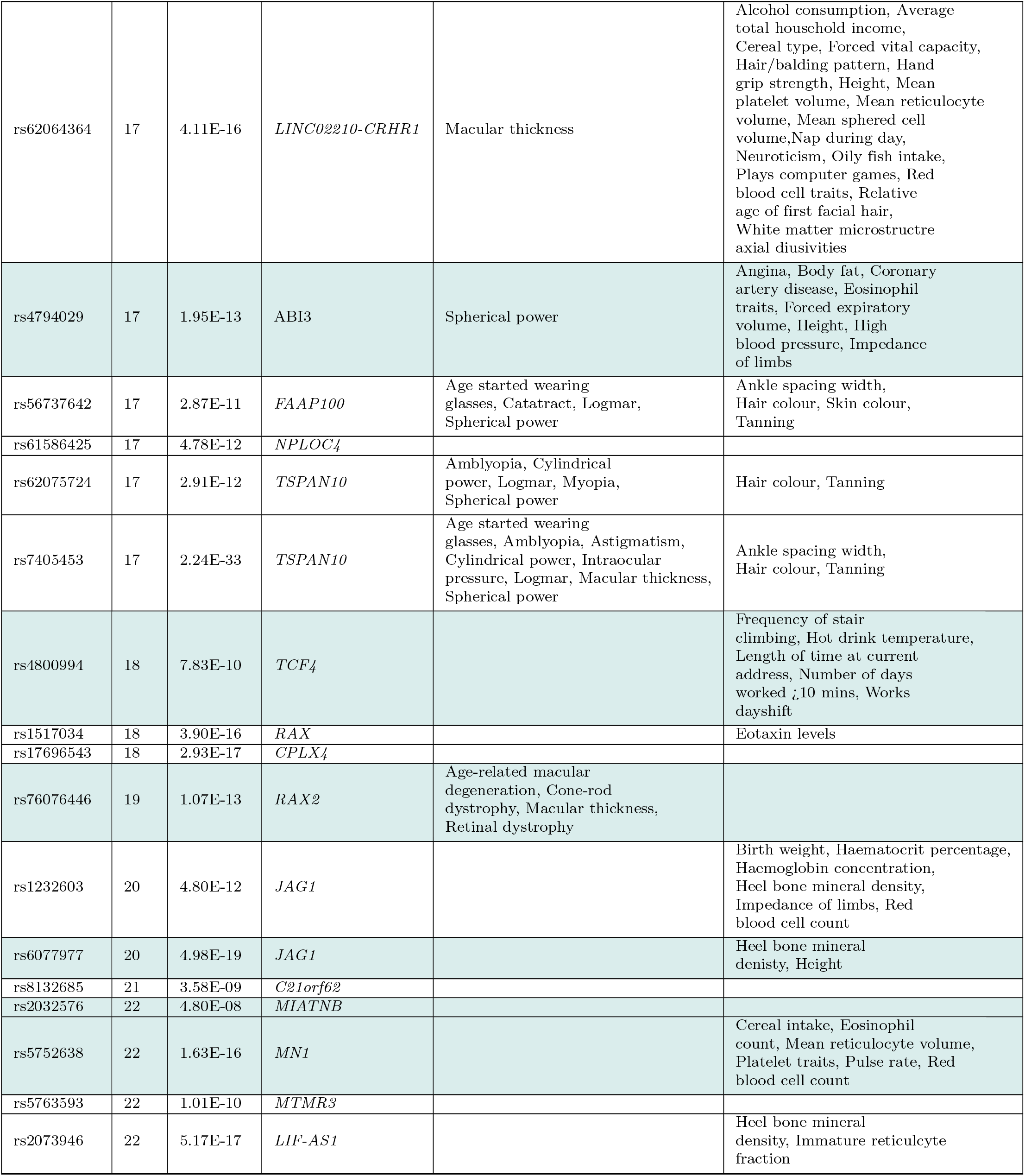
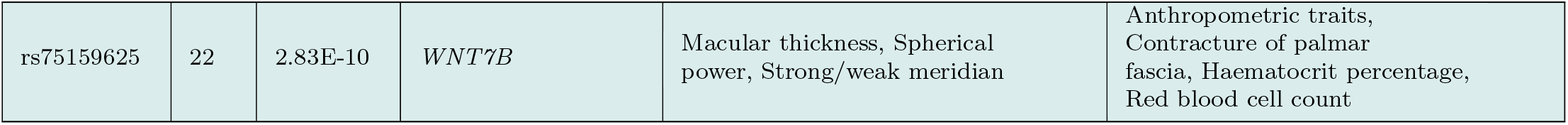
111 SNPs significantly associated (P *<*5 × 10^−8^) with the thickness of one or more of the component photoreceptor cells (PRCs). Each SNP is annotated with the associated gene and any prior associations to ocular or non-ocular phenotypes. Variants considered to be representative of a single locus, examples of allelic heterogeneity, are highlighted in the same colour alternating white and grey. Full results including effect size, effect allele specification and standard error are available in Supplementary Table S4.

**Table S4.** Expanded results of photoreceptor layer thickness genome-wide association studies. Extended results of GWAS of the three photoreceptor cell layers, the outer nuclear layer (ONL), inner segment (IS) and outer segment (OS) following meta-analysis as well as the results selected by meta-analysis using MTAG, labelled MTAG. Each locus found significantly associated following meta-analysis is listed alongside the chromosome (Chr) and location (BP). The reference and alternative alleles are listed alongside the allele frequency (AF) of the alternative allele (A1). The effect size refers to the effect of having an additional copy of the A1 allele and the standard error (SE) is further listed.

| SNP | Chr | BP | A1 | A2 | AF | MTAG effect size | ONL effect size | OS effect size | IS effect size | MTAG p-value | ONL p-value | OS p-value | IS p-value | MTAG SE | ONL SE | OS SE | IS SE |
| --- | --- | --- | --- | --- | --- | --- | --- | --- | --- | --- | --- | --- | --- | --- | --- | --- | --- |
| rs112248193 | 1 | 3718402 | A | G | 0.13 | -0.09 | -0.26 | -0.02 | -0.09 | 2.73E-09 | 5.43E-04 | 0.60 | 2.73E-09 | 0.02 | 0.07 | 0.05 | 0.02 |
| rs2128416 | 1 | 10700448 | C | T | 0.15 | -0.53 | -0.53 | -0.02 | -0.06 | 4.83E-13 | 4.83E-13 | 0.63 | 1.99E-04 | 0.07 | 0.07 | 0.04 | 0.02 |
| rs11587687 | 1 | 110147801 | A | G | 0.44 | 0.18 | -0.08 | 0.18 | 0.01 | 5.02E-09 | 0.12 | 5.02E-09 | 0.30 | 0.03 | 0.05 | 0.03 | 0.01 |
| rs3790612 | 1 | 113084146 | A | G | 0.26 | -0.27 | -0.08 | -0.27 | -0.01 | 2.60E-15 | 0.17 | 2.60E-15 | 0.55 | 0.03 | 0.06 | 0.03 | 0.01 |
| rs72683442 | 1 | 113468825 | C | T | 0.22 | -0.35 | -0.20 | -0.35 | -0.02 | 4.13E-21 | 1.18E-03 | 4.13E-21 | 0.16 | 0.04 | 0.06 | 0.04 | 0.01 |
| rs410895 | 1 | 196898226 | C | T | 0.55 | 0.30 | 0.06 | 0.30 | -0.01 | 8.19E-22 | 0.23 | 8.19E-22 | 0.17 | 0.03 | 0.05 | 0.03 | 0.01 |
| rs6427827 | 1 | 200398387 | A | T | 0.62 | 0.42 | 0.42 | -0.04 | 0.05 | 2.50E-16 | 2.50E-16 | 0.22 | 2.23E-06 | 0.05 | 0.05 | 0.03 | 0.01 |
| rs11577827 | 1 | 202798831 | T | C | 0.53 | 0.37 | 0.37 | 0.07 | 0.05 | 2.77E-13 | 2.77E-13 | 0.03 | 6.56E-06 | 0.05 | 0.05 | 0.03 | 0.01 |
| rs919655 | 1 | 214157972 | A | G | 0.12 | 0.46 | 0.46 | 2.03E-03 | 0.08 | 2.36E-09 | 2.36E-09 | 0.97 | 4.00E-07 | 0.08 | 0.08 | 0.05 | 0.02 |
| rs12140498 | 1 | 222098690 | T | C | 0.20 | 0.37 | 0.37 | 0.08 | 0.05 | 3.14E-09 | 3.14E-09 | 0.04 | 2.89E-05 | 0.06 | 0.06 | 0.04 | 0.01 |
| rs6426584 | 1 | 227374949 | T | A | 0.34 | 0.07 | 0.28 | -0.03 | 0.07 | 7.03E-11 | 1.62E-07 | 0.36 | 7.03E-11 | 0.01 | 0.05 | 0.03 | 0.01 |
| rs7594221 | 2 | 24112724 | A | T | 0.76 | 0.28 | 0.23 | 0.28 | 0.03 | 1.07E-14 | 8.19E-05 | 1.07E-14 | 0.01 | 0.04 | 0.06 | 0.04 | 0.01 |
| rs116350483 | 2 | 145338686 | T | C | 0.02 | -2.11 | -2.11 | -0.16 | -0.33 | 2.10E-26 | 2.10E-26 | 0.19 | 2.86E-15 | 0.20 | 0.20 | 0.12 | 0.04 |
| rs80265589 | 2 | 169027979 | A | T | 0.27 | -0.42 | -0.42 | -0.14 | -0.06 | 1.86E-13 | 1.86E-13 | 2.80E-05 | 4.21E-08 | 0.06 | 0.06 | 0.03 | 0.01 |
| rs28416292 | 2 | 182507470 | A | T | 0.15 | 0.08 | 0.34 | -0.02 | 0.08 | 1.78E-08 | 1.61E-06 | 0.61 | 1.78E-08 | 0.01 | 0.07 | 0.04 | 0.01 |
| rs58172089 | 2 | 198916926 | A | G | 0.17 | 0.23 | -0.07 | 0.23 | -0.03 | 2.74E-08 | 0.32 | 2.74E-08 | 0.03 | 0.04 | 0.07 | 0.04 | 0.01 |
| rs3755152 | 2 | 216822197 | G | A | 0.89 | -0.31 | 0.42 | -0.31 | 0.05 | 4.39E-10 | 5.15E-07 | 4.39E-10 | 1.83E-03 | 0.05 | 0.08 | 0.05 | 0.02 |
| rs148388367 | 2 | 216850944 | A | T | 0.04 | -1.30 | -1.30 | 0.56 | -0.14 | 1.20E-21 | 1.20E-21 | 9.63E-12 | 1.18E-06 | 0.14 | 0.14 | 0.08 | 0.03 |
| rs201030469 | 2 | 218321984 | G | A | 0.14 | -0.42 | -0.42 | -0.01 | -0.07 | 6.58E-09 | 6.58E-09 | 0.82 | 1.30E-05 | 0.07 | 0.07 | 0.04 | 0.02 |
| rs7564805 | 2 | 234228946 | G | A | 0.05 | -0.78 | -0.07 | -0.78 | 0.01 | 6.16E-29 | 0.52 | 6.16E-29 | 0.55 | 0.07 | 0.12 | 0.07 | 0.02 |
| rs34234056 | 3 | 14418444 | G | A | 0.51 | 0.17 | 0.01 | 0.17 | -1.87E-03 | 2.37E-08 | 0.78 | 2.37E-08 | 0.86 | 0.03 | 0.05 | 0.03 | 0.01 |
| rs11129176 | 3 | 25049310 | A | G | 0.28 | 0.32 | 0.32 | 0.02 | 0.04 | 1.53E-08 | 1.53E-08 | 0.60 | 1.31E-04 | 0.06 | 0.06 | 0.03 | 0.01 |
| rs6775323 | 3 | 27721085 | T | G | 0.20 | 0.49 | 0.49 | -0.04 | 0.08 | 5.79E-15 | 5.79E-15 | 0.34 | 4.35E-09 | 0.06 | 0.06 | 0.04 | 0.01 |
| rs62282867 | 3 | 100972078 | A | G | 0.79 | 0.34 | -0.16 | 0.34 | -1.98E-03 | 4.63E-19 | 0.01 | 4.63E-19 | 0.88 | 0.04 | 0.06 | 0.04 | 0.01 |
| rs111163508 | 3 | 129235423 | C | G | 0.16 | -0.39 | 0.23 | -0.39 | 0.03 | 2.28E-20 | 9.22E-04 | 2.28E-20 | 0.03 | 0.04 | 0.07 | 0.04 | 0.01 |
| rs7430585 | 3 | 150112041 | A | G | 0.76 | 0.23 | 0.10 | 0.23 | -0.05 | 1.26E-10 | 0.09 | 1.26E-10 | 2.66E-05 | 0.04 | 0.06 | 0.04 | 0.01 |
| rs115237855 | 5 | 17186336 | A | G | 0.03 | -0.68 | 0.17 | -0.68 | 0.03 | 3.73E-12 | 0.29 | 3.73E-12 | 0.31 | 0.10 | 0.16 | 0.10 | 0.03 |
| rs78303234 | 5 | 17218432 | G | C | 0.12 | -0.26 | -0.10 | -0.26 | 0.01 | 3.32E-08 | 0.21 | 3.32E-08 | 0.58 | 0.05 | 0.08 | 0.05 | 0.02 |
| rs30373 | 5 | 55745334 | G | C | 0.37 | 0.32 | 0.32 | 0.11 | 0.04 | 7.70E-10 | 7.70E-10 | 3.86E-04 | 4.10E-05 | 0.05 | 0.05 | 0.03 | 0.01 |
| rs63338061 | 5 | 71486228 | T | C | 0.36 | 0.07 | 0.31 | -7.24E-05 | 0.07 | 9.81E-11 | 3.71E-09 | 0.99 | 9.81E-11 | 0.01 | 0.05 | 0.03 | 0.01 |
| rs17421627 | 5 | 87847586 | G | T | 0.07 | 1.15 | 1.15 | 0.16 | 0.17 | 3.30E-33 | 3.30E-33 | 0.01 | 4.57E-17 | 0.10 | 0.10 | 0.06 | 0.02 |
| rs62391700 | 5 | 126093395 | C | T | 0.20 | -0.45 | -0.45 | -0.13 | -0.03 | 5.05E-13 | 5.05E-13 | 7.46E-04 | 0.04 | 0.06 | 0.06 | 0.04 | 0.01 |
| rs1109114 | 5 | 148615946 | T | C | 0.43 | 0.37 | 0.37 | -0.04 | 0.07 | 2.65E-13 | 2.65E-13 | 0.23 | 1.28E-10 | 0.05 | 0.05 | 0.03 | 0.01 |
| rs1438692 | 5 | 148659664 | A | G | 0.42 | 0.07 | 0.32 | -0.01 | 0.07 | 1.32E-11 | 3.59E-10 | 0.75 | 1.32E-11 | 0.01 | 0.05 | 0.03 | 0.01 |
| rs6875105 | 5 | 173054917 | C | T | 0.38 | -0.38 | -0.38 | -0.03 | -0.03 | 1.39E-13 | 1.39E-13 | 0.27 | 4.48E-03 | 0.05 | 0.05 | 0.03 | 0.01 |
| rs2326838 | 6 | 6901663 | A | G | 0.36 | -0.23 | -0.07 | -0.23 | -1.50E-03 | 7.86E-13 | 0.18 | 7.86E-13 | 0.89 | 0.03 | 0.05 | 0.03 | 0.01 |
| rs12192672 | 6 | 7229619 | A | G | 0.30 | -0.34 | -0.34 | 0.01 | -0.04 | 8.88E-10 | 8.88E-10 | 0.76 | 1.13E-04 | 0.06 | 0.06 | 0.03 | 0.01 |
| rs17507554 | 6 | 11394287 | A | G | 0.05 | 0.69 | 0.69 | -0.11 | 0.10 | 3.93E-09 | 3.93E-09 | 0.14 | 4.14E-05 | 0.12 | 0.12 | 0.07 | 0.02 |
| rs6923949 | 6 | 35496366 | G | A | 0.82 | 0.23 | 0.36 | 0.23 | 0.04 | 4.22E-09 | 4.95E-08 | 4.22E-09 | 9.96E-04 | 0.04 | 0.07 | 0.04 | 0.01 |
| rs375435 | 6 | 42661404 | C | T | 0.57 | -0.25 | -0.06 | -0.25 | -0.01 | 3.56E-16 | 0.20 | 3.56E-16 | 0.62 | 0.03 | 0.05 | 0.03 | 0.01 |
| rs6910414 | 6 | 56726737 | G | A | 0.18 | 0.42 | 0.42 | -0.16 | -0.01 | 1.87E-10 | 1.87E-10 | 7.37E-05 | 0.67 | 0.07 | 0.07 | 0.04 | 0.01 |
| rs947340 | 6 | 76747944 | C | A | 0.70 | -0.25 | -0.05 | -0.25 | 0.03 | 1.17E-13 | 0.34 | 1.17E-13 | 0.02 | 0.03 | 0.06 | 0.03 | 0.01 |
| rs74526772 | 6 | 106515218 | A | T | 0.04 | -1.25 | -1.25 | -0.01 | -0.18 | 1.13E-19 | 1.13E-19 | 0.90 | 6.24E-10 | 0.14 | 0.14 | 0.08 | 0.03 |
| rs9639276 | 7 | 867033 | T | C | 0.17 | 0.41 | 0.41 | -0.01 | 0.08 | 4.74E-10 | 4.74E-10 | 0.75 | 3.79E-09 | 0.07 | 0.07 | 0.04 | 0.01 |
| rs12531825 | 7 | 8005174 | A | G | 0.12 | 0.45 | 0.45 | 0.09 | 0.08 | 5.94E-09 | 5.94E-09 | 0.07 | 2.34E-06 | 0.08 | 0.08 | 0.05 | 0.02 |
| rs12719025 | 7 | 51100190 | G | A | 0.46 | 0.49 | 0.49 | -3.73E-03 | 0.07 | 6.39E-22 | 6.39E-22 | 0.90 | 1.33E-10 | 0.05 | 0.05 | 0.03 | 0.01 |
| rs111963714 | 7 | 99948655 | G | T | 0.21 | 0.27 | 0.20 | 0.27 | 0.03 | 5.96E-13 | 1.54E-03 | 5.96E-13 | 0.05 | 0.04 | 0.06 | 0.04 | 0.01 |
| rs34926272 | 7 | 129591807 | C | G | 0.03 | -1.05 | -1.05 | -0.15 | -0.18 | 2.03E-11 | 2.03E-11 | 0.11 | 4.44E-08 | 0.16 | 0.16 | 0.10 | 0.03 |
| rs62490856 | 8 | 10469030 | A | G | 0.13 | 0.33 | -0.09 | 0.33 | 0.04 | 5.02E-13 | 0.25 | 5.02E-13 | 0.01 | 0.05 | 0.08 | 0.05 | 0.02 |
| rs61675430 | 8 | 61671071 | A | G | 0.20 | -0.41 | -0.41 | -7.74E-04 | -0.06 | 1.30E-10 | 1.30E-10 | 0.98 | 1.33E-05 | 0.06 | 0.06 | 0.04 | 0.01 |

Table S4. continued
|  |  |  |  |  |  |  |  |  |  |  |  |  |  |  |  |  |  |
| --- | --- | --- | --- | --- | --- | --- | --- | --- | --- | --- | --- | --- | --- | --- | --- | --- | --- |
| rs13263941 | 8 | 109121945 | C | T | 0.26 | 0.86 | 0.86 | 0.15 | 0.12 | 2.07E-50 | 2.07E-50 | 1.72E-05 | 6.09E-24 | 0.06 | 0.06 | 0.04 | 0.01 |
| rs376067714 | 8 | 109141863 | G | A | 0.18 | 0.77 | 0.77 | 0.06 | 0.11 | 2.73E-24 | 2.73E-24 | 0.17 | 4.14E-12 | 0.08 | 0.08 | 0.05 | 0.02 |
| rs9298817 | 9 | 21576591 | C | A | 0.66 | -0.51 | -0.51 | -0.04 | -0.04 | 2.90E-22 | 2.90E-22 | 0.21 | 6.77E-04 | 0.05 | 0.05 | 0.03 | 0.01 |
| rs10781177 | 9 | 76593011 | T | C | 0.42 | -0.29 | -0.29 | -0.01 | -0.05 | 1.49E-08 | 1.49E-08 | 0.64 | 1.12E-05 | 0.05 | 0.05 | 0.03 | 0.01 |
| rs717299 | 9 | 77185933 | G | A | 0.45 | 0.37 | 0.37 | 0.06 | 0.04 | 3.80E-13 | 3.80E-13 | 0.04 | 9.57E-05 | 0.05 | 0.05 | 0.03 | 0.01 |
| rs111245635 | 10 | 48389841 | T | C | 0.02 | 1.26 | 1.26 | 0.30 | 0.23 | 8.41E-10 | 8.41E-10 | 0.02 | 1.27E-07 | 0.21 | 0.21 | 0.12 | 0.04 |
| rs1947075 | 10 | 49741135 | T | C | 0.64 | -0.32 | -0.32 | -0.07 | -0.05 | 7.86E-10 | 7.86E-10 | 0.04 | 3.66E-06 | 0.05 | 0.05 | 0.03 | 0.01 |
| rs7916697 | 10 | 69991853 | G | A | 0.76 | -0.43 | -0.43 | -0.14 | -0.06 | 3.80E-13 | 3.80E-13 | 1.40E-04 | 1.51E-07 | 0.06 | 0.06 | 0.04 | 0.01 |
| rs11200922 | 10 | 85961758 | G | A | 0.46 | 0.33 | 0.08 | 0.33 | -0.03 | 1.62E-26 | 0.12 | 1.62E-26 | 0.01 | 0.03 | 0.05 | 0.03 | 0.01 |
| rs34309160 | 10 | 104034550 | T | C | 0.07 | -0.53 | -0.53 | -0.04 | -0.06 | 3.34E-08 | 3.34E-08 | 0.55 | 0.01 | 0.10 | 0.10 | 0.06 | 0.02 |
| rs17102399 | 10 | 123435963 | A | G | 0.05 | 0.67 | 0.67 | -0.02 | 0.07 | 2.44E-08 | 2.44E-08 | 0.75 | 3.95E-03 | 0.12 | 0.12 | 0.07 | 0.03 |
| rs60401382 | 10 | 124227624 | T | C | 0.23 | -0.30 | 0.07 | -0.30 | 0.03 | 2.43E-16 | 0.27 | 2.43E-16 | 0.01 | 0.04 | 0.06 | 0.04 | 0.01 |
| rs1016934 | 11 | 31720621 | G | A | 0.30 | -0.39 | -0.39 | 0.02 | -0.03 | 2.18E-12 | 2.18E-12 | 0.46 | 0.01 | 0.06 | 0.06 | 0.03 | 0.01 |
| rs618838 | 11 | 66328719 | C | T | 0.55 | 0.06 | 0.19 | 0.03 | 0.06 | 1.79E-09 | 1.49E-04 | 0.28 | 1.79E-09 | 0.01 | 0.05 | 0.03 | 0.01 |
| rs116233906 | 11 | 68968271 | A | C | 0.04 | -0.94 | -0.94 | -0.04 | -0.11 | 9.33E-14 | 9.33E-14 | 0.59 | 3.09E-05 | 0.13 | 0.13 | 0.08 | 0.03 |
| rs10737153 | 11 | 69281829 | C | A | 0.57 | -0.30 | -0.30 | -0.02 | -0.03 | 3.18E-09 | 3.18E-09 | 0.48 | 3.00E-03 | 0.05 | 0.05 | 0.03 | 0.01 |
| rs12574286 | 11 | 76937602 | C | G | 0.21 | -0.29 | 0.01 | -0.29 | 0.01 | 5.77E-15 | 0.93 | 5.77E-15 | 0.41 | 0.04 | 0.06 | 0.04 | 0.01 |
| rs1126809 | 11 | 89017961 | A | G | 0.30 | -0.32 | 0.20 | -0.32 | 0.04 | 2.56E-22 | 2.49E-04 | 2.56E-22 | 1.04E-03 | 0.03 | 0.05 | 0.03 | 0.01 |
| rs6483429 | 11 | 95239787 | T | C | 0.46 | -0.30 | -0.30 | 8.18E-04 | -0.04 | 2.67E-09 | 2.67E-09 | 0.98 | 4.72E-04 | 0.05 | 0.05 | 0.03 | 0.01 |
| rs2080402 | 12 | 345175 | C | T | 0.56 | 0.22 | -0.13 | 0.22 | -0.03 | 2.21E-12 | 0.01 | 2.21E-12 | 3.18E-03 | 0.03 | 0.05 | 0.03 | 0.01 |
| rs3138142 | 12 | 56115585 | T | C | 0.24 | 0.96 | 0.96 | 0.48 | 0.10 | 5.58E-60 | 5.58E-60 | 5.01E-41 | 7.10E-17 | 0.06 | 0.06 | 0.04 | 0.01 |
| rs76629482 | 12 | 96178789 | G | C | 0.18 | -0.53 | -0.53 | -0.06 | -0.07 | 3.38E-16 | 3.38E-16 | 0.151 | 3.69E-07 | 0.07 | 0.07 | 0.04 | 0.01 |
| rs9796234 | 13 | 114323997 | C | T | 0.52 | 0.27 | 0.19 | 0.27 | 0.03 | 4.69E-18 | 1.57E-04 | 4.69E-18 | 3.13E-03 | 0.03 | 0.05 | 0.03 | 0.01 |
| rs28468687 | 14 | 36026342 | A | G | 0.18 | 0.38 | 0.38 | 0.06 | 0.05 | 1.02E-08 | 1.02E-08 | 0.13 | 2.01E-04 | 0.07 | 0.07 | 0.04 | 0.01 |
| rs1254260 | 14 | 60835737 | A | G | 0.29 | -0.20 | 0.25 | -0.20 | -0.03 | 1.18E-09 | 7.86E-06 | 1.18E-09 | 0.01 | 0.03 | 0.06 | 0.03 | 0.01 |
| rs1956524 | 14 | 68800393 | A | G | 0.64 | 0.29 | 0.29 | 0.02 | 0.04 | 2.13E-08 | 2.13E-08 | 0.59 | 1.09E-03 | 0.05 | 0.05 | 0.03 | 0.01 |
| rs10135971 | 14 | 69517494 | A | G | 0.34 | 0.43 | 0.43 | -0.02 | 0.03 | 4.47E-16 | 4.47E-16 | 0.45 | 0.01 | 0.05 | 0.05 | 0.03 | 0.01 |
| rs112145470 | 14 | 74356090 | A | G | 0.03 | 1.34 | 1.34 | 0.09 | 0.14 | 4.81E-16 | 4.81E-16 | 0.40 | 3.64E-05 | 0.17 | 0.17 | 0.10 | 0.03 |
| rs368205955 | 14 | 74497636 | G | T | 0.02 | -3.19 | -3.19 | -0.24 | -0.50 | 4.13E-46 | 4.13E-46 | 0.08 | 1.56E-26 | 0.22 | 0.22 | 0.14 | 0.05 |
| rs12147951 | 14 | 74642451 | C | A | 0.07 | -2.34 | -2.34 | -0.10 | -0.38 | 8.89E-112 | 8.89E-112 | 0.13 | 4.78E-67 | 0.10 | 0.10 | 0.06 | 0.02 |
| rs1972565 | 14 | 74666824 | G | A | 0.82 | 1.91 | 1.91 | 0.11 | 0.30 | 5.23E-191 | 5.23E-191 | 0.01 | 7.95E-106 | 0.06 | 0.06 | 0.04 | 0.01 |
| rs1972564 | 14 | 74666944 | T | C | 0.48 | 1.03 | 1.03 | -0.04 | 0.15 | 2.93E-74 | 2.93E-74 | 0.22 | 1.40E-37 | 0.06 | 0.06 | 0.03 | 0.01 |
| rs118186707 | 14 | 74686575 | A | G | 0.03 | 1.77 | 1.77 | 0.03 | 0.27 | 1.70E-32 | 1.70E-32 | 0.74 | 5.61E-18 | 0.15 | 0.15 | 0.09 | 0.03 |
| rs28488340 | 14 | 74693803 | G | C | 0.40 | -0.59 | -0.59 | -0.05 | -0.08 | 9.02E-31 | 9.02E-31 | 0.14 | 1.07E-13 | 0.05 | 0.05 | 0.03 | 0.01 |
| rs888413 | 14 | 75267396 | T | C | 0.47 | 0.39 | 0.39 | 0.07 | 0.07 | 1.92E-14 | 1.92E-14 | 0.02 | 3.64E-10 | 0.05 | 0.05 | 0.03 | 0.01 |
| rs1800407 | 15 | 28230318 | T | C | 0.08 | 0.57 | -0.03 | 0.57 | -0.02 | 7.58E-25 | 0.70246515 | 7.58E-25 | 0.34 | 0.06 | 0.09 | 0.06 | 0.02 |
| rs1648303 | 15 | 45445692 | G | A | 0.57 | -0.28 | -0.28 | -0.01 | -0.04 | 2.72E-08 | 2.72E-08 | 0.66 | 8.32E-04 | 0.05 | 0.05 | 0.03 | 0.01 |
| rs10083695 | 15 | 53987147 | G | A | 0.49 | -0.36 | -0.36 | -0.06 | -0.06 | 8.00E-13 | 8.00E-13 | 0.03 | 5.67E-09 | 0.05 | 0.05 | 0.03 | 0.01 |
| rs3825991 | 15 | 89761664 | A | C | 0.47 | 0.36 | 0.05 | 0.36 | -0.04 | 2.70E-31 | 0.29 | 2.70E-31 | 1.10E-04 | 0.03 | 0.05 | 0.03 | 0.01 |
| rs1372613 | 15 | 101204835 | T | C | 0.30 | 0.35 | 0.35 | 0.02 | 0.06 | 1.31E-10 | 1.31E-10 | 0.63 | 2.60E-07 | 0.05 | 0.05 | 0.03 | 0.01 |
| rs7206532 | 16 | 80490131 | C | T | 0.52 | 0.30 | 0.30 | 0.02 | 0.03 | 1.86E-09 | 1.86E-09 | 0.56 | 3.06E-03 | 0.05 | 0.05 | 0.03 | 0.01 |
| rs142963458 | 16 | 84561361 | T | C | 0.04 | -0.73 | 0.77 | -0.73 | 0.13 | 3.58E-18 | 3.55E-08 | 3.58E-18 | 4.56E-06 | 0.08 | 0.14 | 0.08 | 0.03 |
| rs1049868 | 16 | 85706633 | C | T | 0.29 | -0.34 | -0.34 | 0.02 | -0.05 | 1.91E-09 | 1.91E-09 | 0.61 | 3.52E-05 | 0.06 | 0.06 | 0.03 | 0.01 |
| rs62064364 | 17 | 43654468 | T | C | 0.22 | 0.50 | 0.50 | 0.02 | 0.07 | 4.11E-16 | 4.11E-16 | 0.65 | 1.71E-07 | 0.06 | 0.06 | 0.04 | 0.01 |
| rs4794029 | 17 | 47280301 | C | T | 0.68 | 0.24 | 0.06 | 0.24 | -0.04 | 1.95E-13 | 0.27 | 1.95E-13 | 1.28E-04 | 0.03 | 0.05 | 0.03 | 0.01 |
| rs56737642 | 17 | 79515509 | G | A | 0.48 | -0.34 | -0.34 | 0.21 | -0.06 | 2.87E-11 | 2.87E-11 | 4.64E-11 | 3.90E-08 | 0.05 | 0.05 | 0.03 | 0.01 |
| rs61586425 | 17 | 79590835 | A | C | 0.09 | -0.48 | -0.14 | -0.48 | -3.22E-03 | 4.78E-12 | 0.22 | 4.78E-12 | 0.89 | 0.07 | 0.11 | 0.07 | 0.02 |
| rs62075724 | 17 | 79611410 | C | T | 0.46 | 0.38 | 0.38 | -0.21 | 0.07 | 2.91E-12 | 2.91E-12 | 1.12E-10 | 1.94E-09 | 0.05 | 0.05 | 0.03 | 0.01 |
| rs7405453 | 17 | 79615572 | G | A | 0.65 | -0.39 | 0.40 | -0.39 | 0.07 | 2.24E-33 | 4.81E-14 | 2.24E-33 | 3.40E-10 | 0.03 | 0.05 | 0.03 | 0.01 |
| rs4800994 | 18 | 53403850 | T | C | 0.19 | 0.08 | 0.31 | -0.12 | 0.08 | 7.83E-10 | 1.33E-06 | 1.88E-03 | 7.83E-10 | 0.01 | 0.06 | 0.04 | 0.01 |
| rs1517034 | 18 | 56937489 | A | G | 0.31 | 0.45 | 0.45 | -1.38E-03 | 0.07 | 3.90E-16 | 3.90E-16 | 0.97 | 9.89E-11 | 0.05 | 0.05 | 0.03 | 0.01 |
| rs17696543 | 18 | 56971398 | T | C | 0.18 | -0.56 | -0.56 | 4.79E-03 | -0.09 | 2.93E-17 | 2.93E-17 | 0.90 | 2.62E-10 | 0.07 | 0.07 | 0.04 | 0.01 |
| rs76076446 | 19 | 3771586 | A | G | 0.02 | -1.25 | -1.25 | 0.14 | -0.15 | 1.07E-13 | 1.07E-13 | 0.19 | 2.19E-05 | 0.17 | 0.17 | 0.10 | 0.04 |
| rs1232603 | 20 | 10612963 | T | C | 0.33 | -0.23 | -4.45E-03 | -0.23 | 0.01 | 4.80E-12 | 0.93 | 4.80E-12 | 0.56 | 0.03 | 0.05 | 0.03 | 0.01 |
| rs6077977 | 20 | 10930708 | G | A | 0.49 | 0.27 | -0.09 | 0.27 | -0.01 | 4.98E-19 | 0.08 | 4.98E-19 | 0.28 | 0.03 | 0.05 | 0.03 | 0.01 |
| rs8132685 | 21 | 34220618 | T | C | 0.52 | -0.30 | -0.30 | -0.02 | -0.04 | 3.58E-09 | 3.58E-09 | 0.54 | 1.01E-04 | 0.05 | 0.05 | 0.03 | 0.01 |
| rs2032576 | 22 | 27089655 | T | G | 0.56 | -0.28 | -0.28 | 0.03 | -0.04 | 4.80E-08 | 4.80E-08 | 0.41 | 1.19E-04 | 0.05 | 0.05 | 0.03 | 0.01 |
| rs5752638 | 22 | 28188203 | C | T | 0.22 | -0.51 | -0.51 | -0.09 | -0.07 | 1.63E-16 | 1.63E-16 | 0.02 | 2.89E-08 | 0.06 | 0.06 | 0.04 | 0.01 |
| rs5763593 | 22 | 30302238 | C | T | 0.63 | 0.34 | 0.34 | -0.01 | 0.06 | 1.01E-10 | 1.01E-10 | 0.74 | 3.12E-09 | 0.05 | 0.05 | 0.03 | 0.01 |
| rs2073946 | 22 | 30619599 | A | G | 0.31 | 0.46 | 0.46 | 0.07 | 0.08 | 5.17E-17 | 5.17E-17 | 0.04 | 2.82E-11 | 0.06 | 0.06 | 0.03 | 0.01 |
| rs75159625 | 22 | 46377008 | G | T | 0.31 | 0.34 | 0.34 | 0.05 | 0.05 | 2.83E-10 | 2.83E-10 | 0.15 | 5.33E-05 | 0.05 | 0.05 | 0.03 | 0.01 |

**Fig S1.**
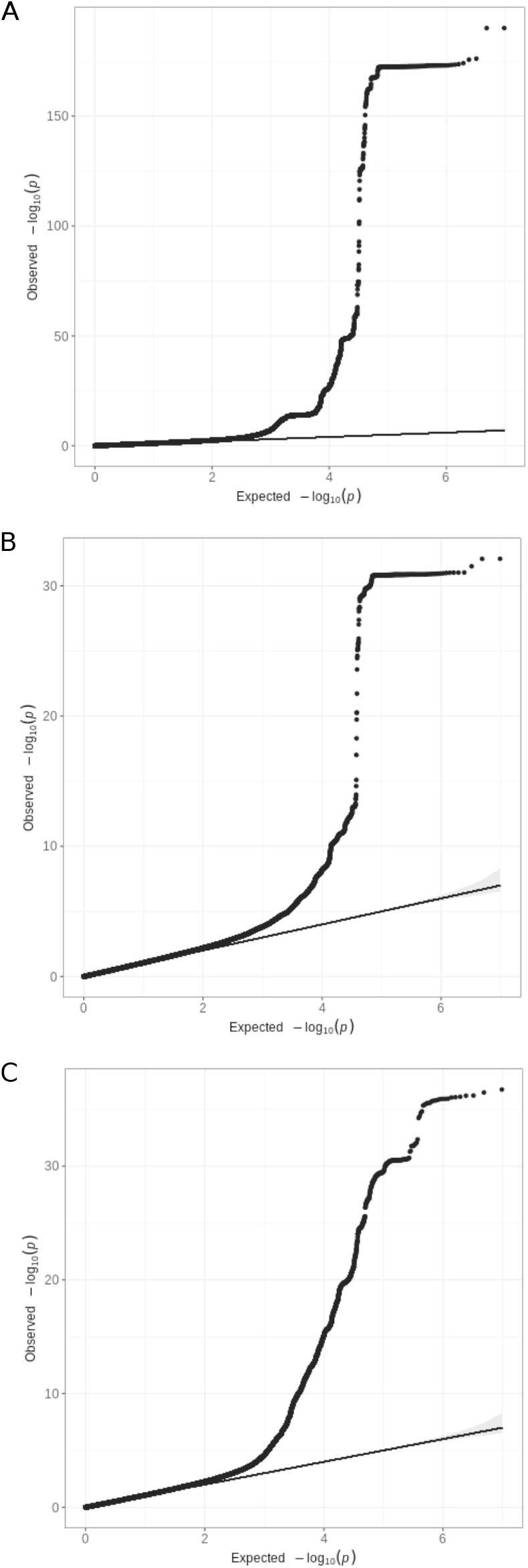
Quantile-quantile plots of outer retinal thickness genome-wide association studies. (A) The quantile-quantile plot (qq-plot) for the GWAS of ONL thickness prior to meta-analysis (Lambda GC = 1.15, Intercept = 1.04, Ratio = 0.13). (B) The qq-plot for the GWAS of IS thickness prior to meta-analysis (Lambda GC = 1.07, Intercept = 1, Ratio *<*0). (C) The qq-plot for the GWAS of OS thickness prior to meta-analysis (Lambda GC = 1.09, Intercept = 1.02, Ratio = 0.15).

**Table S5.** Selected rare monogenic ocular disease genes found to be associated with photoreceptor layer thicknesses in the present study. Several genes are involved in bringing about or shaping the photoreceptors’ electrical response to light (*RHO, GNAT2, SAG, GRK1*) or in recycling of the light-sensitive chromophore (*ABCA4, RLBP1, RBP3, RDH5*). Others are involved in maintaining outer retinal structure or in retinal and eye development. Superscript letters denote the relevant analyses in which the association was found: ^a^GWAS; ^b^differential association with ONL across concentric fields; ^c^differential association with IS across concentric fields; ^d^differential association with OS across concentric fields; ^e^exome analysis.

| Gene | Location | Encoded Protein and Function | Mendelian disease | Mode of inheritance |
| --- | --- | --- | --- | --- |
| *Genes associated with rare monogenic retinal diseases* |  |  |  |  |
| *CDHR1*<sup>a,c</sup> | 10q23.1 | Cadherin-related family member 1 (maintains rod and cone outer segment structure) | Cone-rod dystrophy; macular dystrophy | Recessive |
| *CERKL*<sup>a</sup> | 2q31.3 | Ceramide kinase-like protein (role in viability of cells with membranes rich in sphingolipids) | Rod-cone dystrophy; cone-rod dystrophy; widespread retinal degeneration | Recessive |
| *GNAT2*<sup>a</sup> | 1p13.3 | Cone-specific transducin (part of the phototransduction cascade in cone photoreceptors) | Achromatopsia (non-functioning cone photoreceptors) | Recessive |
| *GRK1*<sup>a</sup> | 13q34 | G-protein receptor kinase 1, rhodopsin kinase (phosphorylates activated rhodopsin to permit shut-off of the light response) | Oguchi disease, retinitis pigmentosa | Recessive |
| *IMPG1*<sup>a</sup> | 6q14.1 | Interphotoreceptor matrix proteoglycan 1 (role in retinal cell adhesion/ interphotoreceptor matrix) | Retinitis pigmentosa; vitelliform macular dystrophy | Dominant and recessive |
| *IMPG2*<sup>a</sup> | 3q12.3 | Interphotoreceptor matrix proteoglycan 2 (role in retinal cell adhesion/ interphotoreceptor matrix) | Retinitis pigmentosa; vitelliform macular dystrophy | Dominant and recessive |
| *JAG1*<sup>a</sup> | 20p12.2 | Jagged protein 1 (ligand for Notch proteins; function in cell-cell signalling) | Alagille syndrome | Dominant |
| *PRPH2*<sup>a</sup> | 6p21.1 | Peripherin-2 (important for rod and cone outer segment disc structure) | Macular pattern dystrophy; retinitis pigmentosa | Dominant and recessive |
| *RAX2*<sup>a,c</sup> | 19p13.3 | Retina and anterior neural fold homeobox 2 transcription factor | Retinitis pigmentosa; cone-rod dystrophy | Dominant and recessive |
| *RBP3*<sup>a</sup> | 10q11.22 | Retinol binding protein 3 (role in retinoid recycling) | Retinal dystrophy associated with high myopia | Recessive |
| *RDH5*<sup>a,b,d</sup> | 14q24.1 | Retinol dehydrogenase 5 (role in retinoid recycling) | Fundus albipunctatus, retinitis pigmentosa | Recessive |
| *RHO*<sup>a</sup> | 3q22.1 | Rhodopsin (forms visual pigment in rods) | Retinitis pigmentosa | Dominant and recessive |
| *RLBP1*<sup>a</sup> | 15q26.1 | Retinaldehyde-binding protein 1 (role in retinoid recycling) | Bothnia dystrophy, retinitis pigmentosa | Recessive |
| *RP1L1*<sup>a</sup> | 15q26.1 | Retinitis pigmentosa 1-like protein 1 (role in photoreceptor connecting cilia) | Occult macular dystrophy; retinitis pigmentosa | Dominant and recessive |

**Table S5. continued**
|  |  |  |  |  |
| --- | --- | --- | --- | --- |
| *SAG*<sup>a</sup> | 2q37.1 | S-antigen, arrestin (binds activated rhodopsin to permit shut-off of the light response) | Oguchi disease, retinitis pigmentosa | Recessive and dominant |
| *TULP1*<sup>a</sup> | 6p21.31 | Tubby-like protein 1 (role in rhodopsin transport within photoreceptors) | Retinitis pigmentosa; Leber congenital amaurosis | Recessive |
| *CFH*<sup>d</sup> | 1q31.3 | Complement factor H (regulates alternative complement pathway) | Early onset macular drusen | Dominant |
| *MERTK*<sup>d</sup> | 2q13 | c-mer protooncogene receptor tyrosine kinase (loss of function leads to defective phagocytosis of photoreceptor outer segments by RPE) | Retinitis pigmentosa; early onset retinal dystrophy | Recessive |
| *ABCA4*<sup>e</sup> | 1p22.1 | ATP-binding cassette transporter A4 (functions as a flippase for N-retinylidene-phosphatidylethanolamine; aids clearing of all-trans retinaldehyde) | Stargardt disease macular dystrophy; cone-rod dystrophy | Recessive |
| *NR2E3*<sup>e</sup> | 15q23 | Nuclear receptor subfamily 2 group E3 (transcription factor, role in rod photoreceptor development; loss of function leads to excess of S-cones) | Enhanced S-cone syndrome, Goldmann-Faver syndrome, retinitis pigmentosa | Recessive and dominant |
| *MYO7A*<sup>e</sup> | 11q13.5 | Myosin VIIA (motor protein functions in transport of opsin in photoreceptors and melanosomes in RPE) | Type 1 Usher syndrome (retinitis pigmentosa and deafness) | Recessive |
| ***Genes associated with oculocutaneous albinism (and foveal hypoplasia)*** |  |  |  |  |
| *OCA2*<sup>a,e</sup> | 15q12-15q13.1 | Oculocutaneous albinism 2 melanosomal transmembrane protein (protein regulates pH of melanosomes) | Oculocutaneous albinism | Recessive |
| *TYR*<sup>a</sup> | 11q14.3 | Tyrosinase (role in melanin synthesis) | Oculocutaneous albinism | Recessive |
| ***Genes associated with rare monogenic ocular maldevelopment (including microphthalmia)*** |  |  |  |  |
| *VSX2*<sup>a,b</sup> | 14q24.3 | Visual system homeobox 2 (role in eye development) | Microphthalmia | Recessive |
| *PITX2*<sup>e</sup> | 4q25 | Paired-like homeodomain transcription factor 2 (role in eye development) | Anterior segment dysgenesis; microphthalmia | Dominant |
| *ATOH7*<sup>a</sup> | 10q21.3 | Atonal basic helix-loop-helix transcription factor 7 (role in eye development) | Persistent primary hypoplastic vitreous; ocular maldevelopment, including microphthalmia | Recessive |
| *CHD7*<sup>a</sup> | 8q12.2 | Chromodomain helicase DNA-binding protein 7 (role in control of neurocrest gene expression and in regulation of cell motility) | CHARGE syndrome | Dominant |
| *RARB*<sup>a</sup> | 3p24.2 | Retinoic acid receptor beta (role in eye and other organ development) | Microphthalmia, syndromic | Dominant and recessive |
| *RAX*<sup>a</sup> | 18q21.32 | Retina and anterior neural fold homeobox (role in eye development) | Microphthalmia | Recessive |

**Fig S2.**
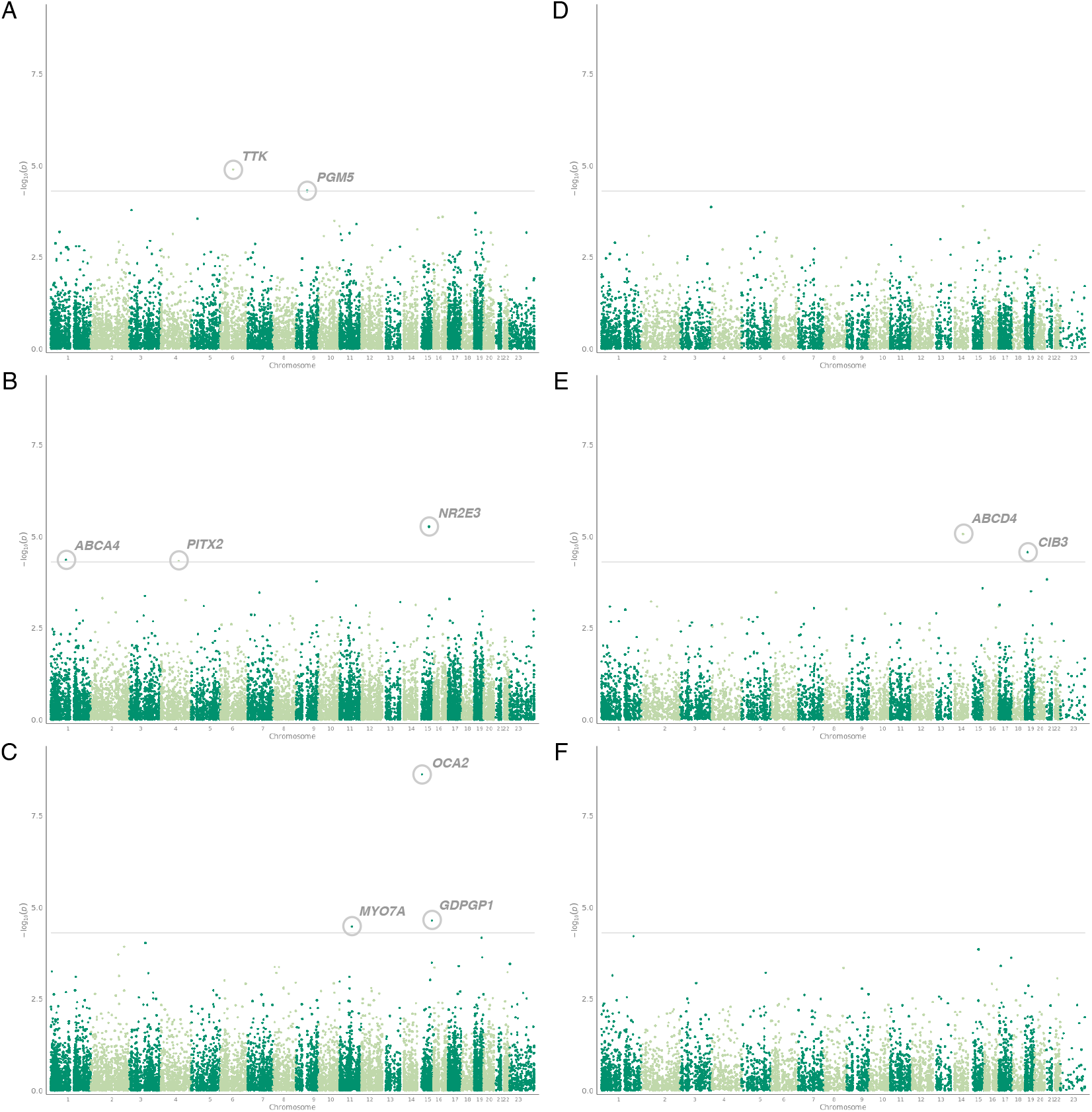
Exome-wide loss of function burden analysis study. Manhattan plot of p-values resulting from exome wide loss of function burden testing analysis for the thickness of each of the retinal layers (A) ONL, missense model, (B) IS, missense model, (C) OS, missense model, (D) ONL, loss of function model, (E) IS, loss of function model, (F) IS, loss of function model. Variants are considered significantly associated if they reach a a p-value threshold of P *<*5 × 10^−5^.

**Table S6.**
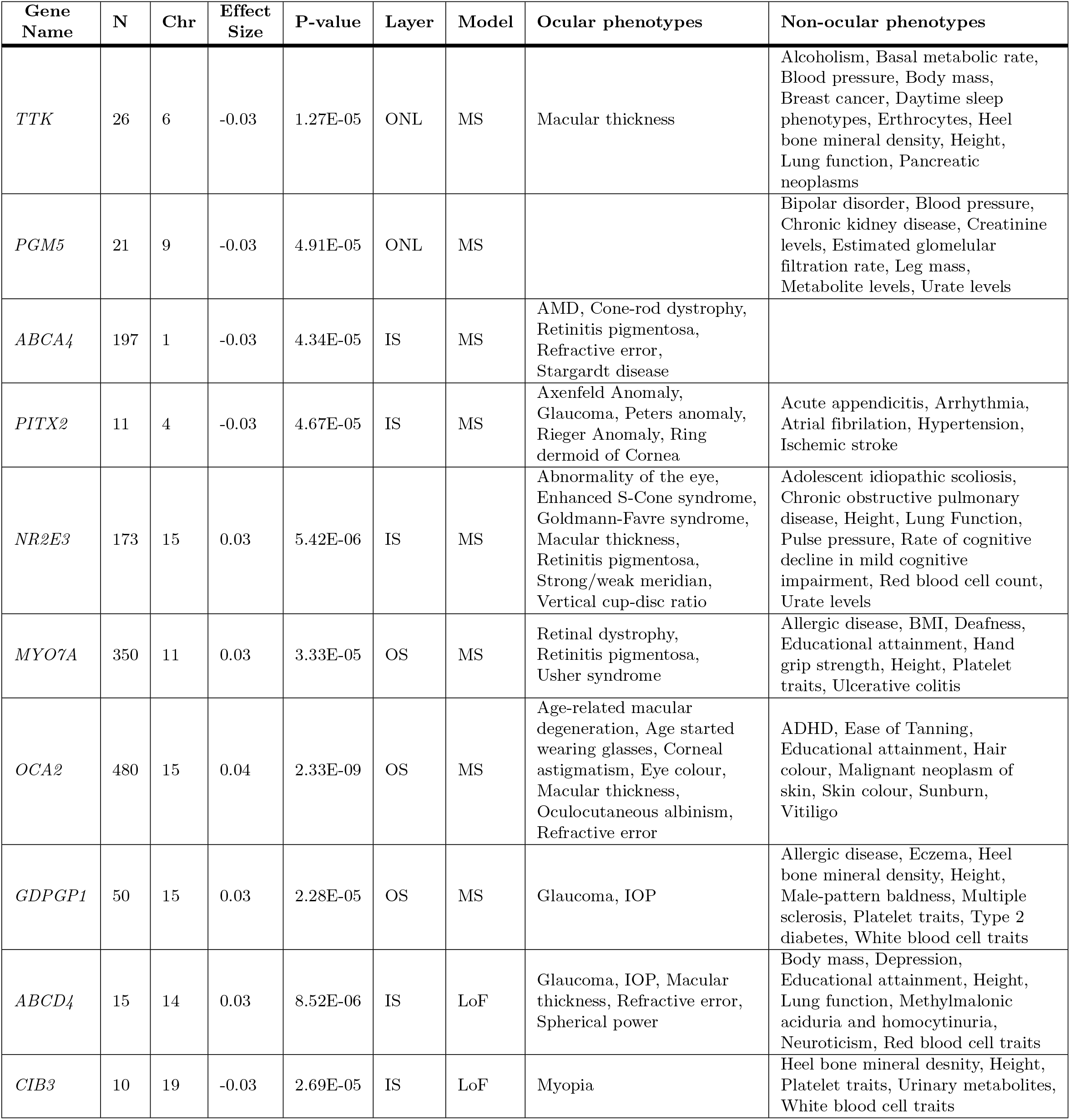
List of genes associated with the thickness of the photoreceptor cell layers following exome analysis. List of genes associated with one of the three photoreceptor layers, the outer nulcear layer (ONL), inner segment (IS) and outer segment (OS) following gene burden testing. A significance threshold of P *<*5 × 10^−5^ is used. For each gene the gene name and chromosome is listed. Each gene is also annotated with any prior associations to ocular and non-ocular phenotypes. The number of individuals with the homozygous loss of function allele is listed as N. Two modes were tested, a missense (MS) model and a loss of function (LoF) model.

**Table S7.**
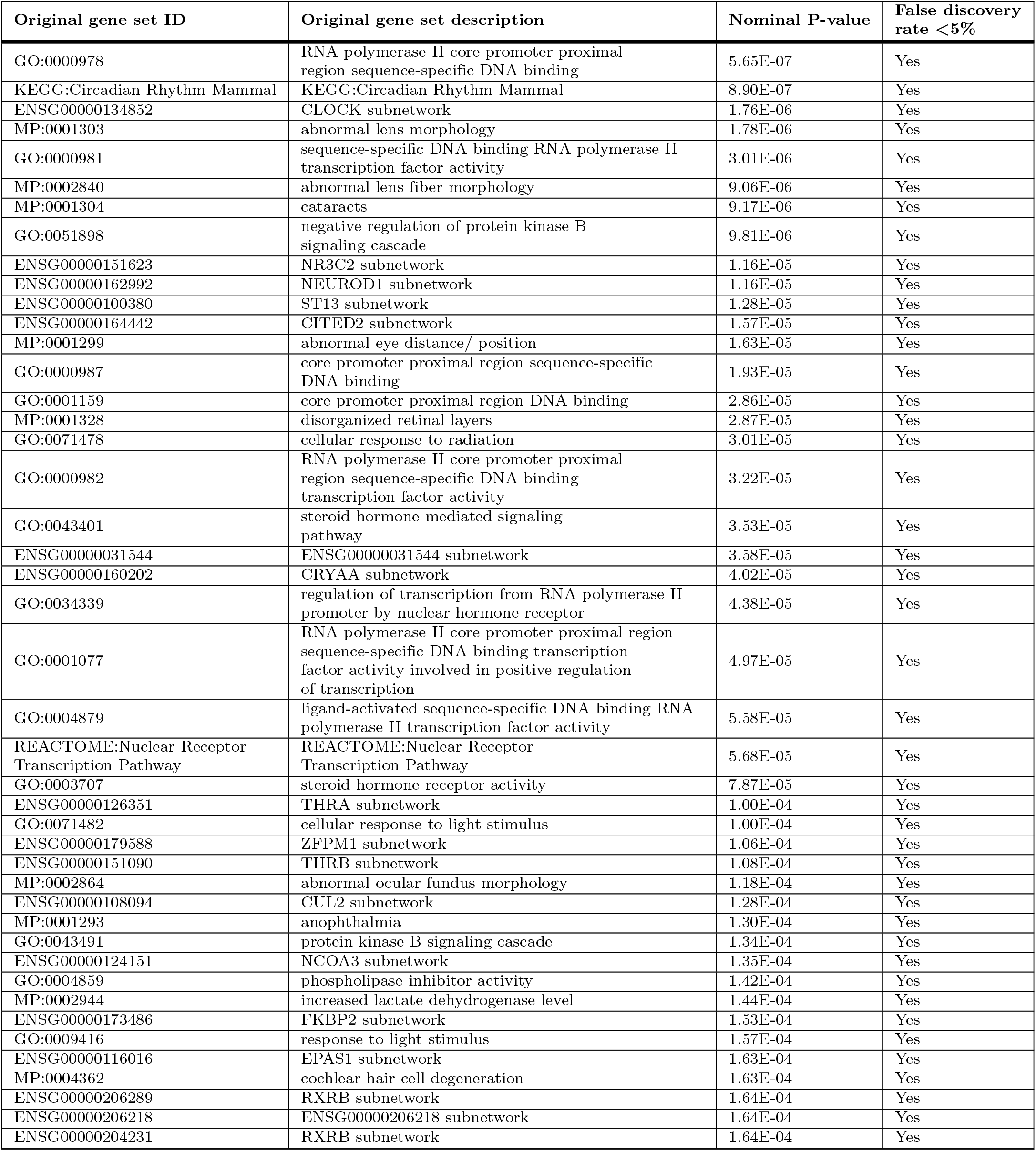
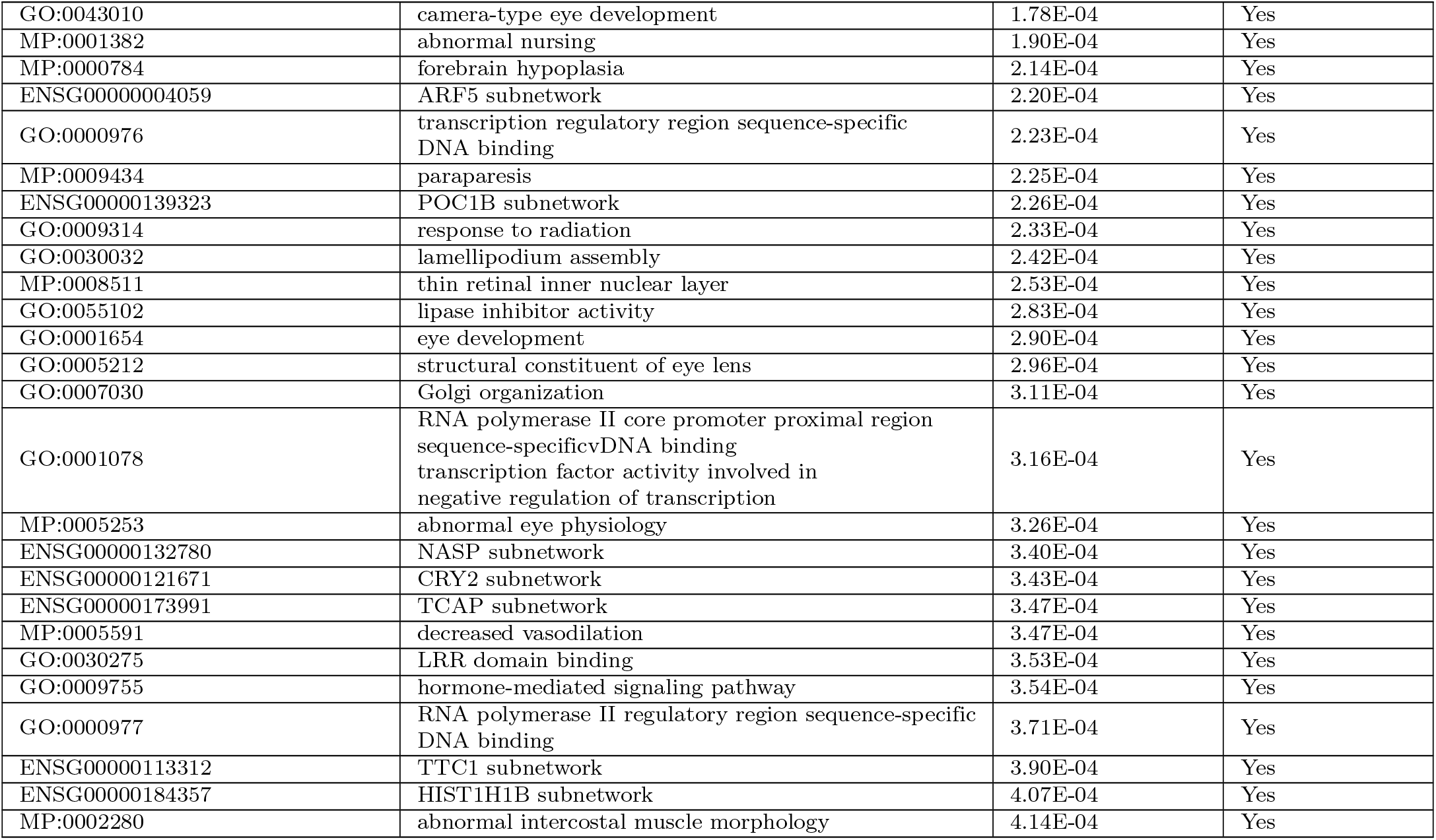
Geneset enrichment analysis. The results of geneset enrichment analysis using DEPICT applied to the GWAS results of the meta analysed PRC layers. All genesets which were significantly enriched following correction for false discovery rate are listed below.

**Table S8.** Tissue enrichment analysis. The results of tissue enrichment analysis using DEPICT applied to the GWAS results of the meta analysed PRC layers. All Tissues which were nominally significantly enriched are listed below.

| MeSH term | MeSH first level term | MeSH second level term | Nominal P value | False discovery rate <5% |
| --- | --- | --- | --- | --- |
| A11.436 | Epithelial Cells | Cells | 1.17E-04 | Yes |
| A09.371 | Eye | Sense Organs | 8.02E-04 | Yes |
| A09.371.729 | Retina | Sense Organs | 1.64E-03 | Yes |
| A10.615 | Membranes | Tissues | 4.41E-03 | No |
| A10.615.550 | Mucous Membrane | Tissues | 0.01 | No |
| A10.615.550.599 | Mouth Mucosa | Tissues | 0.02 | No |
| A03.734.414 | Islets of Langerhans | Digestive System | 0.03 | No |
| A14.549 | Mouth | Stomatognathic System | 0.04 | No |
| A10.272 | Epithelium | Tissues | 0.04 | No |
| A10.615.550.760 | Respiratory Mucosa | Tissues | 0.05 | No |
| A04.531.520 | Nasal Mucosa | Respiratory System | 0.05 | No |
| A09.531 | Nose | Sense Organs | 0.05 | No |

**Fig S3.**
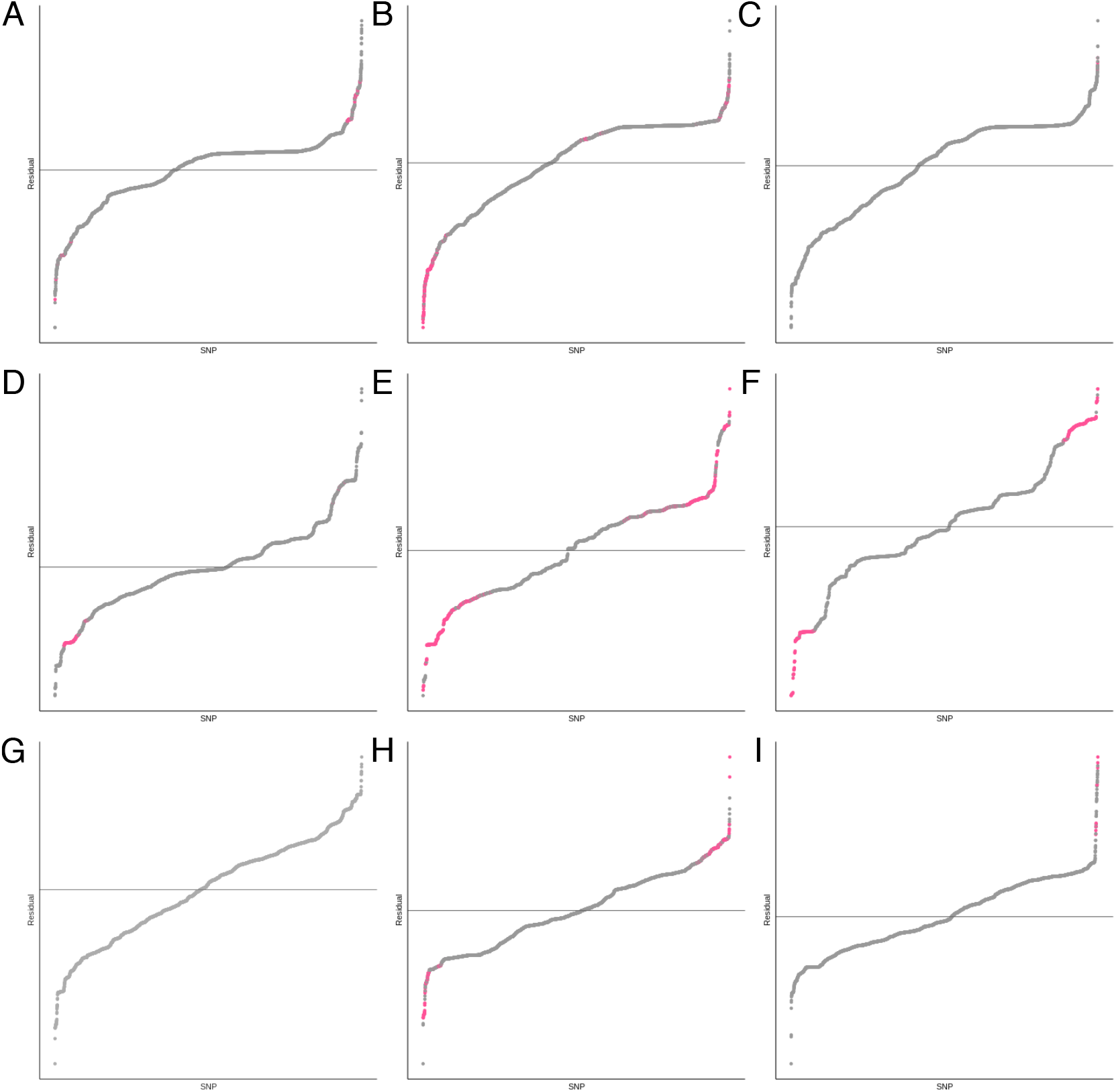
Comparison of the genetic effect on the thickness of concentric retinal fields. Plots depicting the residuals from models comparing the effect size of SNPs on the thickness of each retinal varinats in different concentric retinal fields. Variants with significantly different effect sizes on the two different areas tested, as determined by a z-score, are highlighted in pink. (A) ONL, fovea compared to intermediate; (B) ONL, fovea compared to peripheral; (C) ONL, intermediate compared to peripheral; (D) IS, fovea compared to intermediate; (E) IS, fovea compared to peripheral; (F) IS, intermediate compared to peripheral; (G) OS, fovea compared to intermediate; (H) OS, fovea compared to peripheral; (I) OS, intermediate compared to peripheral.

**Table S9.** List of SNPs differentially affecting the thickness of the ONL at the three concentric fields of the macula. List of SNPs with a significant z-score describing the differential effect on the ONL thickness at the foveal (F), intermediate (I) and peripheral (P) fields. The field (F1 or F2) and corresponding effect size from GWAS of thickness in each field are listed alongside the p-value of the comparative z-score. Each genetic variant is also annotated with associated gene and any ocular and non-ocular phenotypes previously associated with it. The different concentric comparisons are separated by bold horizontal lines.

| SNP | Chr | F1 | F2 | F1 effect size | F2 effect size | P-value | Associated gene | Ocular phenotypes | General phenotypes |
| --- | --- | --- | --- | --- | --- | --- | --- | --- | --- |
| rs11708067 | 3 | F | I | 0.69 | 0.10 | 1.00E-07 | *ADCY5* | Refractive error | Birth length, Birth weight, Cholesterol levels, Chronotype, Chronic kidney disease, Fasting blood glucose, Forced expiratory volume, Heel bone mineral density, Height, Type 2 diabetes |
| rs75757892 | 6 | F | I | -1.12 | -0.55 | 3.75E-06 | *RREB1* | AMD, Cup-to-disc ratio, Refractive error | Birth weight, Blood glucose levels, Chronic kidney disease, Heel bone mineral density, Height, Hematocrit, Hemoglobin concentration, Impedance of body, Microcephaly, Mouth ulcers, Multiple sclerosis, Parkinsons, Red blood cell count, Self-reported math ability, Stroke, Type 2 diabetes, White blood cell count |
| rs45569432 | 14 | F | I | -1.35 | -2.17 | 3.27E-08 | *VSX2* | Macular thickness, Refractive error | Cancer of endocrine glands, Chronotype, Red blood cell traits |
| rs1769287 | 1 | F | P | -0.59 | -0.05 | 6.90E-08 | *SLC1A7* |  | Amyotrophic lateral sclerosis, Duration of fitness test, Height, Hippocampal atrophy, Lupus |
| rs6701735 | 1 | F | P | 0.67 | -0.07 | 5.14E-07 | *MIR29B2CHG* |  | Pulse rate, White blood cell count |
| rs11125573 | 2 | F | P | 0.54 | 0.08 | 2.27E-06 | *CCDC88A* |  | Appendicular lean mass |
| rs11720108 | 3 | F | P | 0.71 | -0.04 | 9.67E-13 | *ADCY5* | Refractive error | Birth weight, Chronic kidney disease, Chronotype, Forced expiratory volume, Heel bone mineral density, Sex hormone-binding globulin levels, Statin medication, Type 2 diabetes |
| rs55914544 | 5 | F | P | 0.65 | -0.04 | 2.53E-08 | *WWC1* |  | Blood pressure, BMI, Body fat, Cerebellum cortex volume, Epithelial ovarian cancer |

Table S9. continued
|  |  |  |  |  |  |  |  |  |  |
| --- | --- | --- | --- | --- | --- | --- | --- | --- | --- |
| rs75757892 | 6 | F | P | -1.12 | -0.20 | 1.84E-15 | *RREB1* | AMD, Cup-to-disc ratio, Refractive error | Birth weight, Blood glucose levels, Chronic kidney disease, Heel bone mineral density, Height, Hematocrit, Hemoglobin concentration, Impedance of body, Microcephaly, Mouth ulcers, Multiple sclerosis, Parkinsons, Red blood cell count, Self-reported math ability, Stroke, Type 2 diabetes, White blood cell count |
| 6:150016812_GA_G | 6 | F | P | 0.61 | 0.13 | 1.54E-06 |  |  |  |
| rs746173658 | 8 | F | P | 0.22 | 0.93 | 4.85E-12 |  |  |  |
| rs1337805 | 9 | F | P | -0.52 | 0.06 | 2.07E-07 | *PTPRD* | Myopia | ADHD, Anxiety, Asthma, Blood pressure, Cancer, Cholesterol, Coronary disease, Depression, Diabetes, Epilepsy, Forced expiratory volume, Heel bone mineral density, Height, Menarche |
| rs11594394 | 10 | F | P | 0.61 | 0.04 | 1.45E-08 | *FRMPD2* | Age started wearing glasses, Myopia, Refractive error, Spherical power |  |
| rs12225226 | 11 | F | P | 0.20 | -0.45 | 2.33E-07 | *LINC01488* |  | BMI, Breast cancer, Cardiomyopathy, Height, Monocyte count, White blood cell count |
| rs3138142 | 12 | F | P | 1.37 | 0.81 | 1.03E-07 | *RDH5* | Age started wearing glasses, Cataract, Early AMD, Fundus albipunctatus, Hypermetropia, Macular thickness, Myopia, Pigmentary retinal dystrophy, Refractive error, Retinal dystrophy, Retinitis pigmentosa, Spherical power |  |
| rs3921811 | 12 | F | P | 0.57 | -0.02 | 7.37E-10 | *PLEKHA5* |  | Time spent outdoors in winter |
| rs1493529 | 13 | F | P | -0.48 | 0.07 | 1.05E-08 | *ENOX1* |  | Arteries, Blood pressure, Depression, End-stage coagulation, Glucose, Mental health disorders, Prostate tumour |
| rs1972565 | 14 | F | P | 1.27 | 1.94 | 8.59E-09 | *VSX2* | Macular thickness, Refractive error | Blood pressure, Chronotype, Coronary artery disease |
| rs3138142 | 12 | I | P | 1.28 | 0.81 | 1.31E-07 | *RDH5* | Age started wearing glasses, Cataract, Early AMD, Fundus albipunctatus, Hypermetropia, Macular thickness, Myopia, Pigmentary retinal dystrophy, Refractive error, Retinal dystrophy, Retinitis pigmentosa, Spherical power |  |

**Table S10.** List of SNPs differentially affecting the thickness of the IS at the three concentric fields of the macula. List of SNPs with a significant z-score describing the differential effect on the IS thickness at the foveal (F), intermediate (I) and peripheral (P) fields. The field (F1 or F2) and corresponding effect size from GWAS of thickness in each field are listed alongside the p-value of the comparative z-score. Each genetic variant is also annotated with associated gene and any ocular and non-ocular phenotypes previously associated with it. The different concentric comparisons are separated by bold horizontal lines.

| SNP | Chr | F1 | F2 | F1 effect size | F2 effect size | P value | Associated gene | Ocular phenotypes | General phenotypes |
| --- | --- | --- | --- | --- | --- | --- | --- | --- | --- |
| rs4721061 | 7 | F | I | -0.23 | -0.09 | 3.40E-11 | *TMEM106B* |  | Coronary artery disease, Dementia, Depression, Height, Irritability, Mood swings, Neuroticism |
| rs55798570 | 10 | F | I | 0.13 | 1.61E-03 | 2.06E-06 | *CDHR1* | Cone-rod dystrophy, Refractive error, Retinitis pigmentosa |  |
| rs61773269 | 1 | F | P | -0.23 | -0.03 | 1.43E-06 | *LRRC8D* |  |  |
| rs7430585 | 3 | F | P | -0.26 | -0.05 | 1.27E-18 | *TSC22D2* | Age started wearing glasses | Blood pressure, Brain morphology, Cholesterol, Platelet count, Pulse pressure, Type 2 diabetes |
| rs59515506 | 4 | F | P | -0.34 | -0.01 | 7.15E-09 | *RUFY3* |  | Cardiovascular disease, Hippocampus, Parkinson disease |
| rs4131080 | 5 | F | P | -0.12 | -0.02 | 1.95E-06 | *HNRNPH1* |  | Neutrophil count, White blood cell count |
| 5:71646122_AATTT_A | 5 | F | P | -0.03 | 0.08 | 4.20E-07 |  |  |  |
| 6:31806799_CA_C | 6 | F | P | -0.13 | 0.01 | 3.33E-08 |  |  |  |
| rs13237518 | 7 | F | P | -0.23 | -0.02 | 1.77E-26 | *TMEM106B* |  | Coronary artery disease, Dementia, Depression, Height, Irritability, Mood swings, Neuroticism, Type 2 diabetes |
| rs141629142 | 9 | F | P | 0.40 | 0.06 | 9.15E-06 | *RORB* | Advanced AMD, Macular thickness, Myopia, Refractive error | Alcohol consumption, Blood pressure, BMI, Cholesterol, Chronotype, Creatinine, Epilepsy, Erythrocytes, Glomerular filtration rate, Glucose, Heart failure, Height |
| rs1409396 | 10 | F | P | 0.13 | 0.02 | 7.63E-09 | *PIP4K2A* |  | Frequency of walking for pleasure, Height, Platelet distribution width |
| rs55798570 | 10 | F | P | 0.13 | -0.06 | 1.12E-13 | *CDHR1* | Cone-rod dystrophy, Refractive error, Retinitis pigmentosa |  |
| rs200916002 | 14 | F | P | -0.07 | -0.21 | 1.46E-06 | *BBOF1* |  |  |
| rs55634267 | 17 | F | P | 0.20 | 0.05 | 1.08E-11 | *GNGT2* | Refractive error, Spherical power | Angina, Asthma, Chronic lower respiratory disease, Coronary artery disease, Eosinophil counts, Hypertension, Impedance of limbs, Metabolite errors, Prostate cancer |

Table S10. continued
|  |  |  |  |  |  |  |  |  |  |
| --- | --- | --- | --- | --- | --- | --- | --- | --- | --- |
| rs850526 | 17 | F | P | -0.16 | -0.04 | 7.56E-08 | *GNGT2* | Spherical power, Refractive error | Appendicular lean mass, Asthma, Chronic lower respiratory disease, Metabolite levels, Primary sclerosing cholangitis, Prostate cancer |
| rs76076446 | 19 | F | P | 0.31 | -0.09 | 1.76E-09 | *RAX2* | AMD, Cone-rod dystrophy, Macular thickness, Retinal dystrophy |  |
| rs2236665 | 21 | F | P | 0.16 | 0.01 | 1.53E-13 | *RRP1B* | Refractive error, Spherical power | Blood pressure, Coronary artery disease, Haemoglobin traits, High blood pressure, Hypertension, Red blood cell traits |
| rs1041451 | 21 | F | P | -0.14 | -0.01 | 2.74E-10 | *C2CD2* |  |  |
| rs6414375 | 3 | I | P | -0.18 | -0.06 | 9.14E-10 | *TSC22D2* | Age started wearing glasses, Glaucoma, IOP, Macular thickness, Refractive error | Blood pressure, Brain morphology, Cholesterol, Platelet count, Pulse pressure, Type 2 diabetes |
| rs13171669 | 5 | I | P | -0.01 | 0.07 | 3.06E-06 | *AFAP1L1* | Macular thickness | Blood pressure, BMI, Forced expiratory volume, Glomerular filtration, Hearing difficulties, Hypertension, Socioeconomic factors |
| rs11974335 | 7 | I | P | -0.09 | -0.01 | 1.15E-05 | *TMEM106B* |  | Dementia, Depression, Height, Impedance of body, Irritability, Mood swings, Neuroticism |
| 21:45115055_CT_C | 21 | I | P | 0.11 | 0.01 | 8.27E-08 |  |  |  |

**Table S11.**
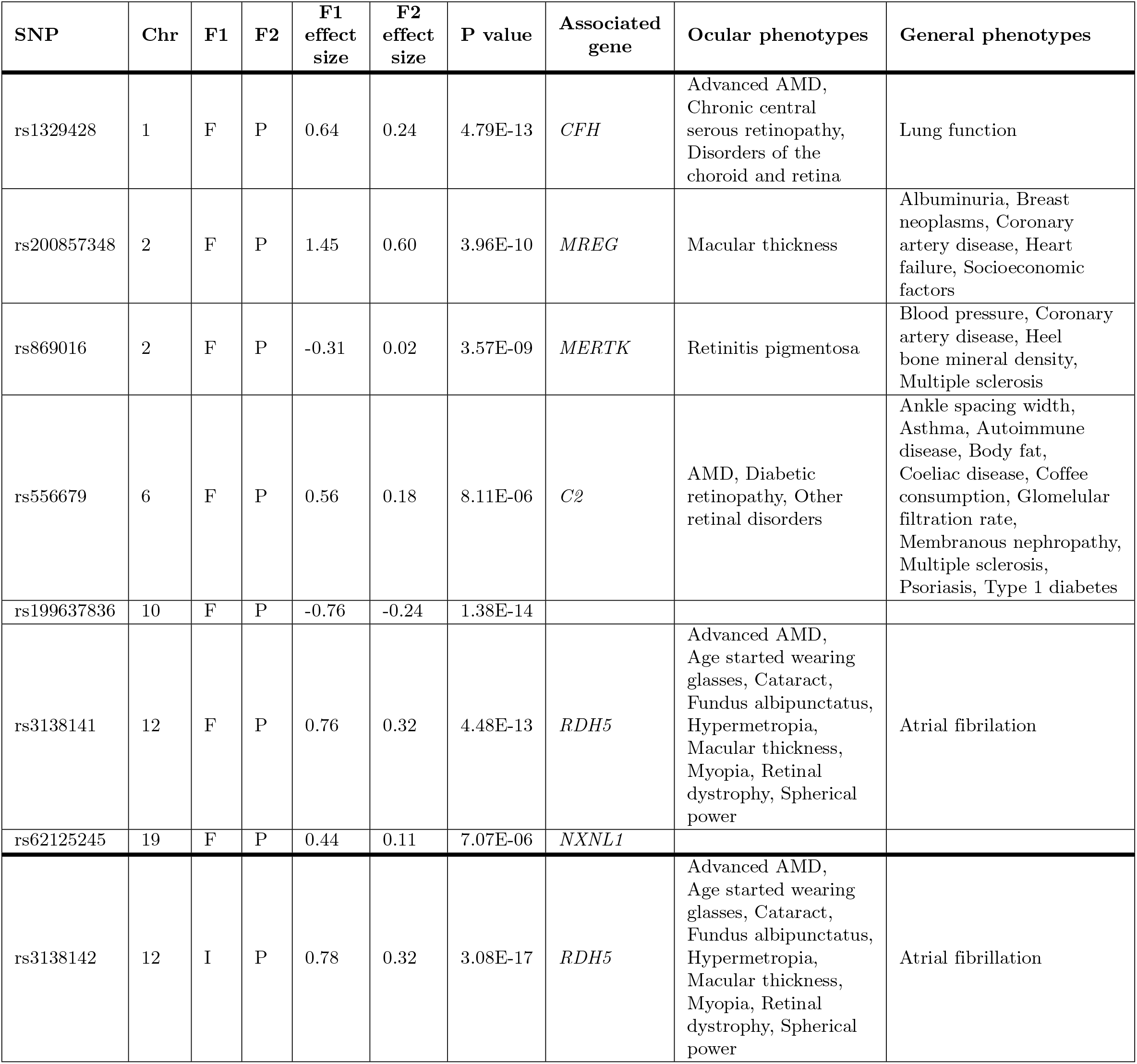
List of SNPs differentially affecting the thickness of the OS at the three concentric fields of the macula. List of SNPs with a significant z-score describing the differential effect on the OS thickness at the foveal (F), intermediate (I) and peripheral (P) fields. The field (F1 or F2) and corresponding effect size from GWAS of thickness in each field are listed alongside the Bonferroni adjusted p-value of the comparative z-score. Each genetic variant is also annotated with associated gene and any ocular and non-ocular phenotypes previously associated it. The different concentric comparisons are separated by bold horizontal lines.

**Fig S4.**
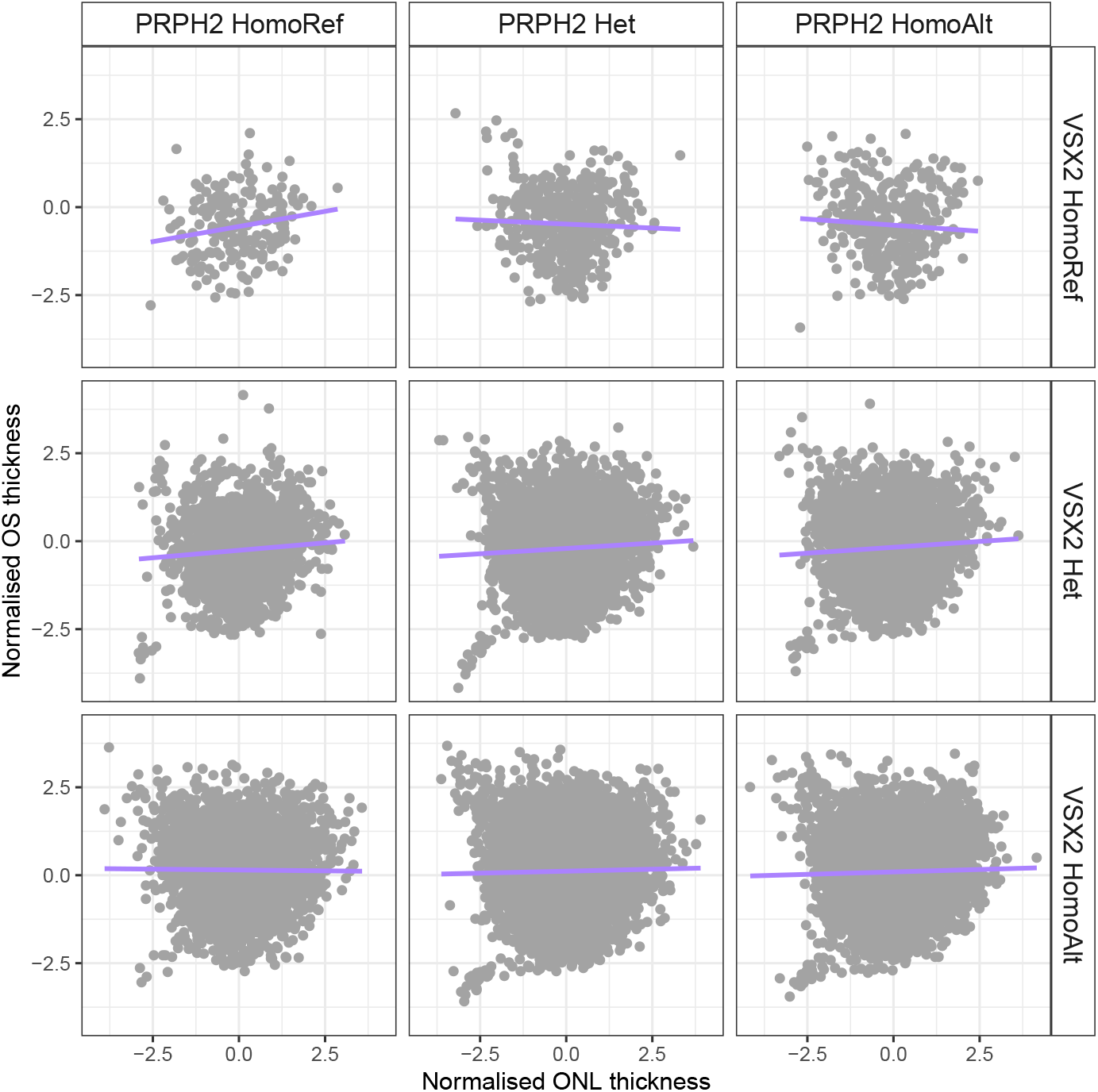
Interaction between genetic variants in *VSX2* and *PRPH2*. Comparison of normalised outer segment (OS) thickness and outer nuclear layer thickness (ONL) in population subsetted by their genotype at *VSX2* (rs1972565) and *PRPH2* (rs375435).

